# Multifractal behavior of SARS-CoV2 COVID-19 pandemic spread, case of: Algeria, Russia, USA and Italy

**DOI:** 10.1101/2020.09.16.20196188

**Authors:** Sid-Ali Ouadfeul

## Abstract

Here, the multifractal behavior SARS-CoV2 COVID-19 pandemic daily and death cases is investigated through the so-called Wavelet Transform Modulus Maxima lines (WTMM) method, data of daily and death cases available via the World Health Organization (WHO) database of Algeria, Russia, USA and Italy is analyzed. The obtained results show the multifractal behavior of the COVID-19 pandemic spread data with different spectra of singularities.

## Introduction

Severe Acute Respiratory Syndrome Coronavirus SARS-CoV2 named also novel coronavirus is a member of coronavidae family. The first human cases of COVID-19, the disease caused by the novel coronavirus causing COVID-19 illness, subsequently named SARS-CoV-2 were first reported by officials in Wuhan City, China, in December 2019. Retrospective investigations by Chinese authorities have identified human cases with onset of symptoms in early December 2019 (WHOa, 2020). COVID-19 can spread from person to person, the possible modes of transmission including contact, droplet, airborne, fomite, fecal-oral, bloodborne, mother-to-child, and animal-to-human transmission. Many mathematical models of COVID-19 spread have been proposed in literature, Berozzi et al (2020) proposed three models of COVID-19 spread which are: the Exponential Growth, the Self-Exciting Branching Process and the Compartmental Models. Iovorra et al (2020) suggested Mathematical modeling of the spread of the coronavirus disease 2019 (COVID-19) taking into account the undetected infections. The case of China was studied. In this paper, the multifractal behavior of COVD-19 pandemic spread investigated through the Wavelet Transform Modulus Maxima lines (WTMM) analysis of daily and death cases. The paper is organized as organized as follows, in section 2 we describe the continuous wavelet transform as a mathematical microscope which is well adapted to scanning the singularities of fractal function. Section 3 is devoted the Takagi model description, while the section 4 is devoted the application of the WTMM method to daily and death cases to investigate the mulifractal behavior. The case of Algeria, Russia, USA and Italy is studied. We end the paper by a conclusion.

### Wavelet Transform Modulus Maxima lines (WTMM)

The wavelet transform modulus lines (WTMM) is a multifractal introduced by Frish and Parisi (1985) revisited by the continuous wavelet transform introduced Grossmann and Morlet in 1985. The WTMM method was first introduced by Malat and Huang in 1992 and used for image processing.

The continuous wavelet transform is a decomposition of a given signal S(t)into a dilated and translated wavelets 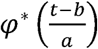 obtained form a mother φ(t) that must have n vanishing moments;

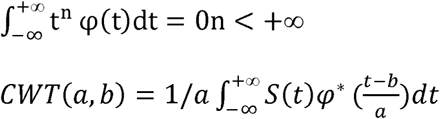

Where *a* ∈ *R*^* +^, *b* ∈ *R*

The first step of WTMM method is to calculate the continuous wavelet transform (CWT) of a given signal and the modulus of CWT, the next step is maxima of the continuous wavelet transform. Determination of local maxima is performed using the computation of the first and second derivative of the wavelet coefficients. CWT *(a,b)* admits a maximum at point *b*_0_ if it satisfies the following two conditions:

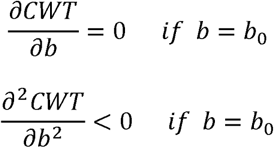

The function of partition *Z*(*q, a*) is a summation of the modulus of the CWT at local maxima *b*_i_ with a *q* power ∈ *R* :

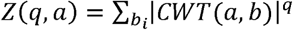

The spectrum of exponents *τ* (*q*) is related the function of partition *Z* (*q, a*)by:

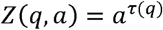

So the spectrum of exponents *τ* (*q*) is obtained by a simple linear regression of log (*Z*(*q, a*)) versus log (*q*)

The spectrum of singularities is the Legendre transform of the spectrum of singularizes:

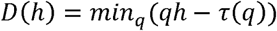

### Takagi model

One the popular Pandemic models is the Takagi curve, construct the curve with the aid of the function *θ:* R → R which measures the distance from every point *x* to its closest integer:

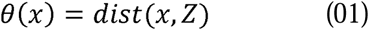

We compute the graph of an approximation of the Takagi function for an *n* great enough in figure 1 using equation 01. This graph is a classical example of a fractal.

**Figure 1.**
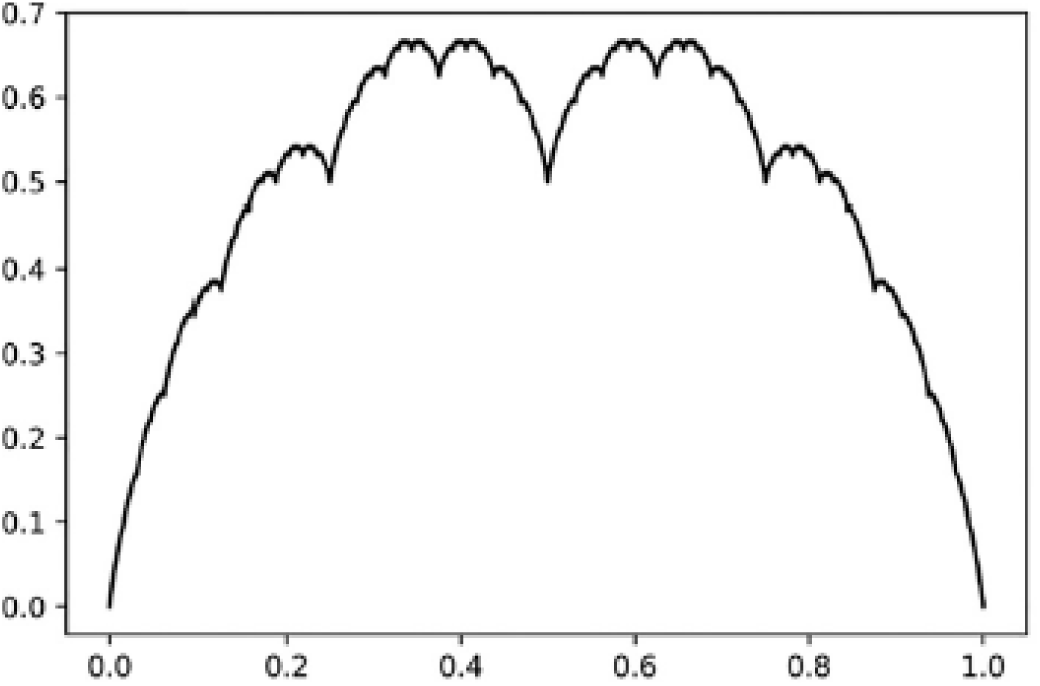
Graph of the Takagi function (Pecurar and Necula, 2020)

Modifying the form of this function as 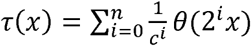 for every *x* ∈ [0,1] with *c* varying between (−2, 2) we obtain new functions. The graph in figure 2 is obtained for computing the function τ*(*x*) for 100 linearly spaced points in the interval [0,1], with *c* randomly chosen in (−2, 2) at each iteration. The C code used is listed below:

**Figure 2.**
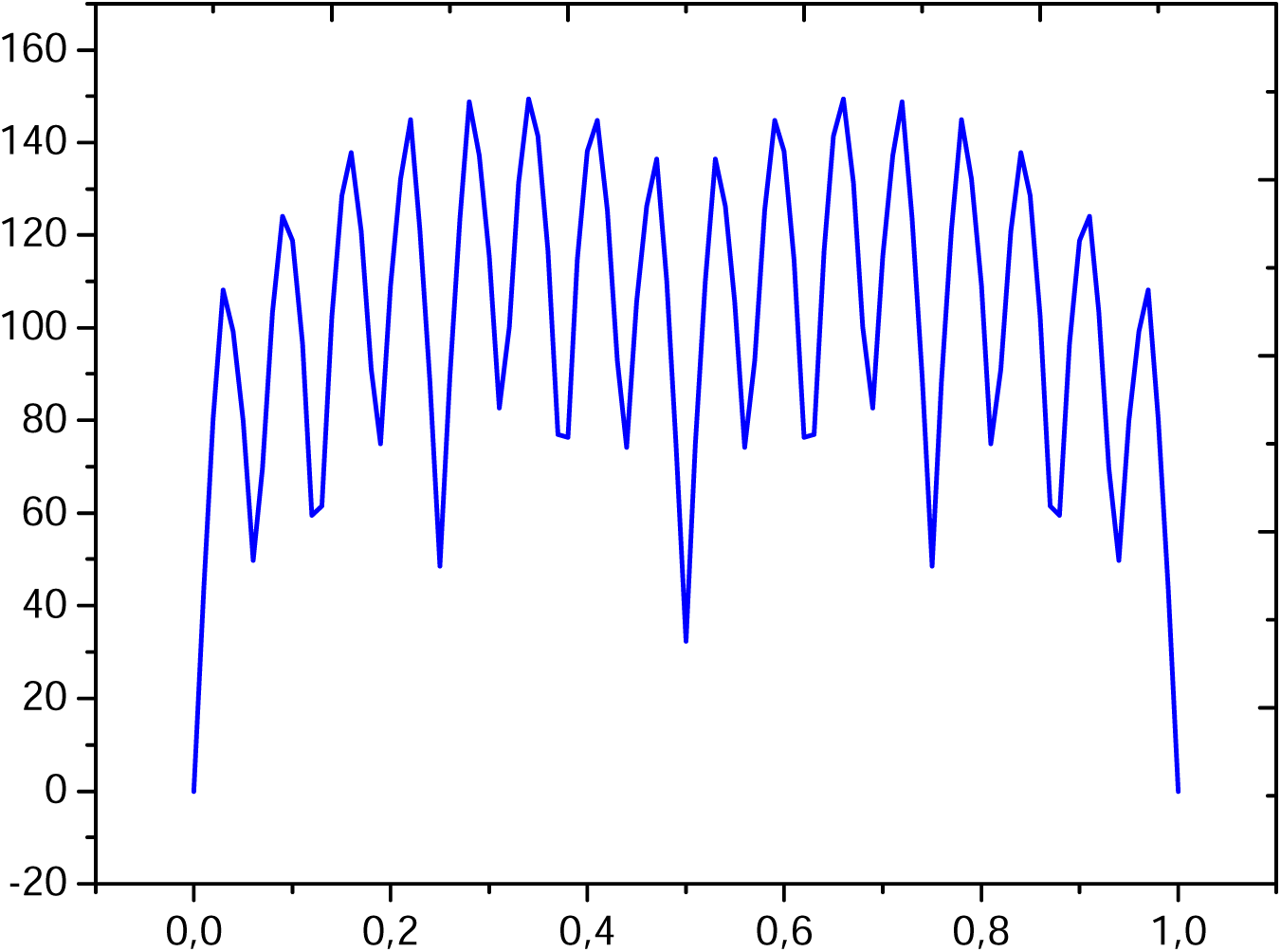
Graph of the randomized modified Takagi function.

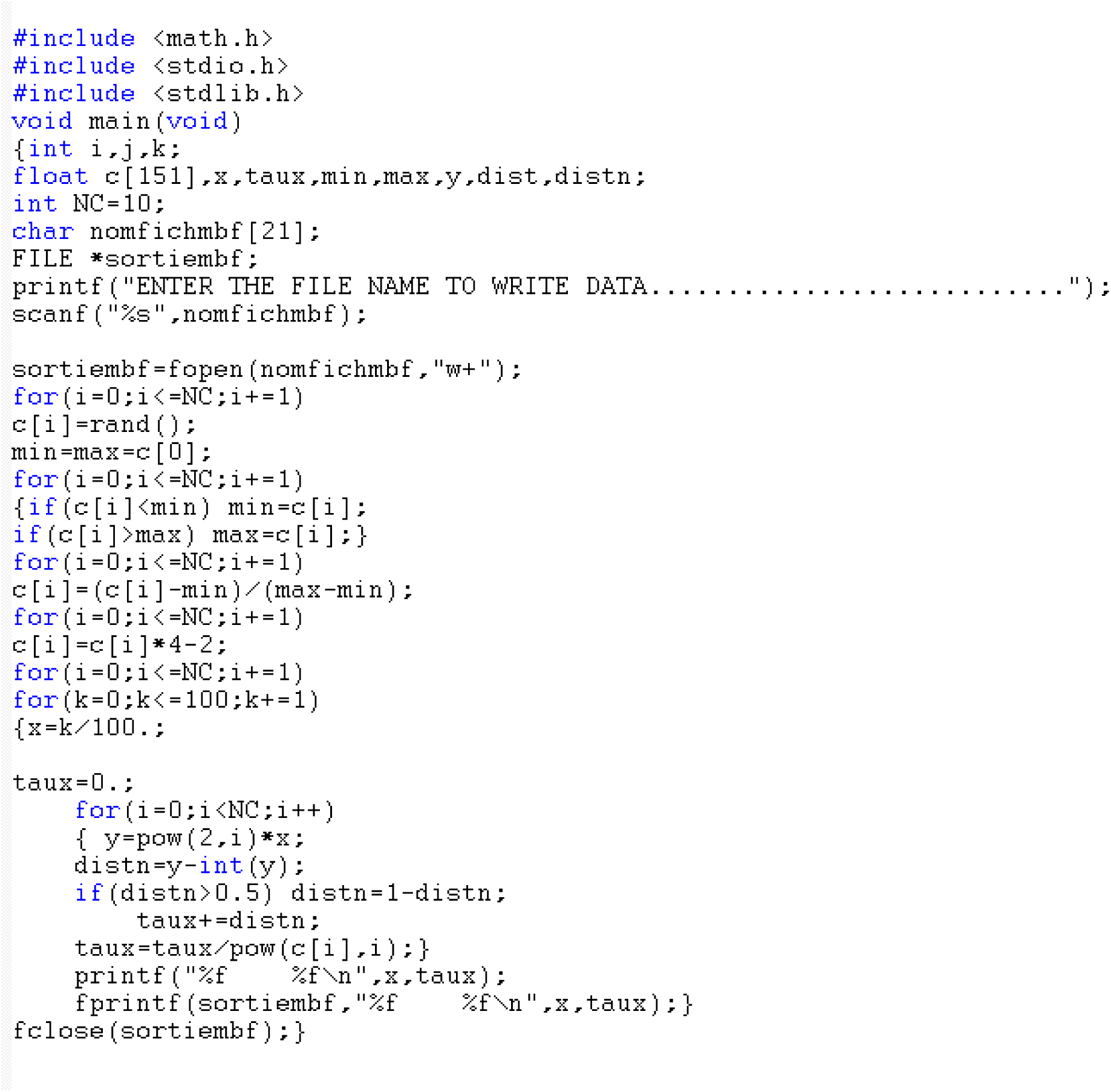

### Case of Algeria

The first case to be analyzed is the daily cases for Algeria, data form Health World Organization (HWO) COVID-19 dashboard are used, figure 3.a shows the graph of the daily cases, the date of first case observed is 25^th^of February 2020, the number of days since the first case is 192.

**Figure 3.a.**
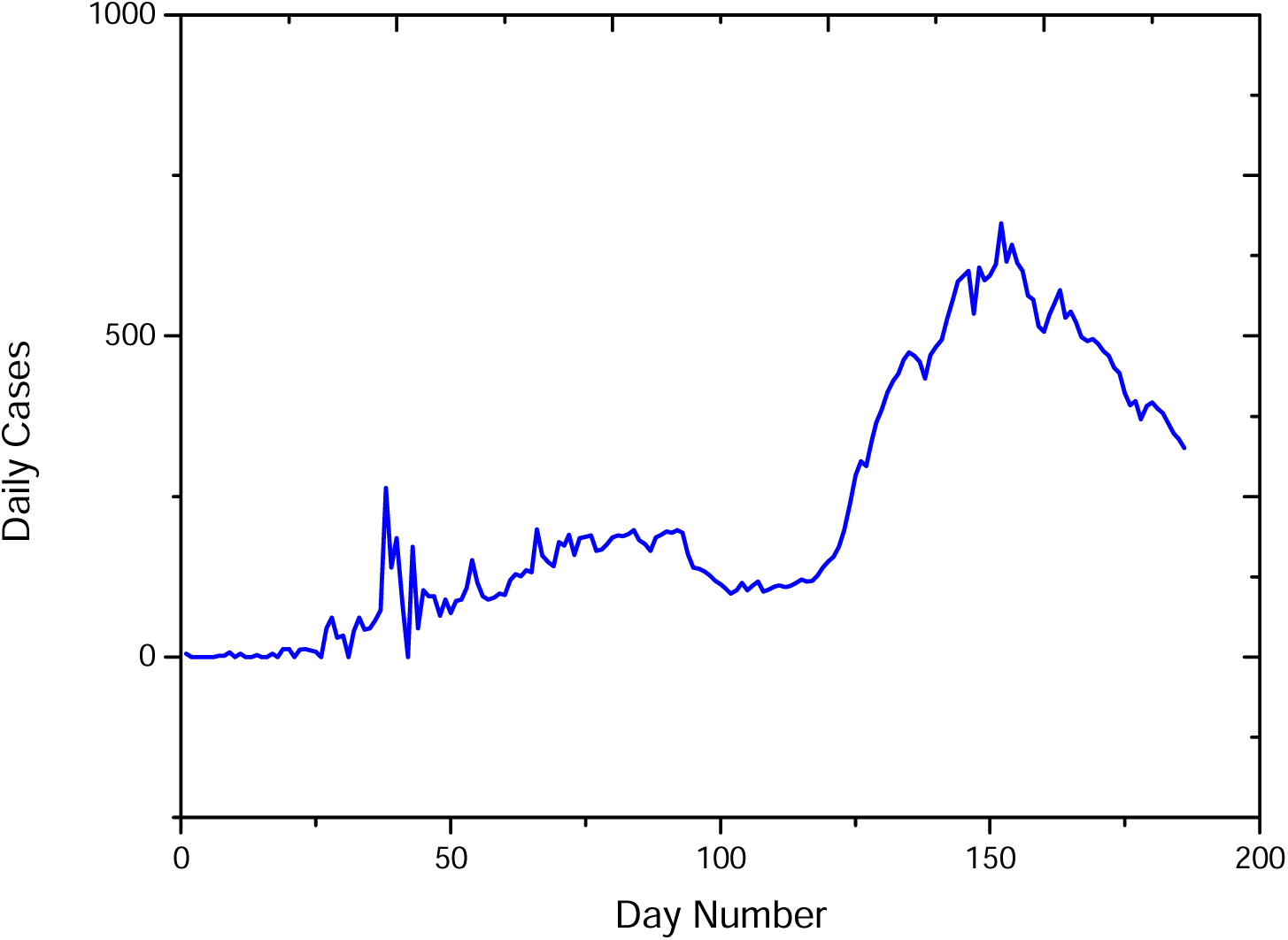
Daily cases in Algeria

Figure 3.b shows the modulus of the continuous wavelet transform in the (time-frequency) domain, analyzing wavelet is the Complex Morlet. Figures 3.c and 3.d show the spectra of exponents and singularities, we observe that the spectrum of exponents has not a linear shape and the spectrum of singularities is not concentrated in one point, which confirm the multifractal behavior of the COVID-19 daily cases in Algeria. High Holder exponents are dominant in the spectrum of singularities which confirm the absence of the high frequency components in the daily cases signal, it means a progressive increase/decrease in the daily cases.

**Figure 3.b.**
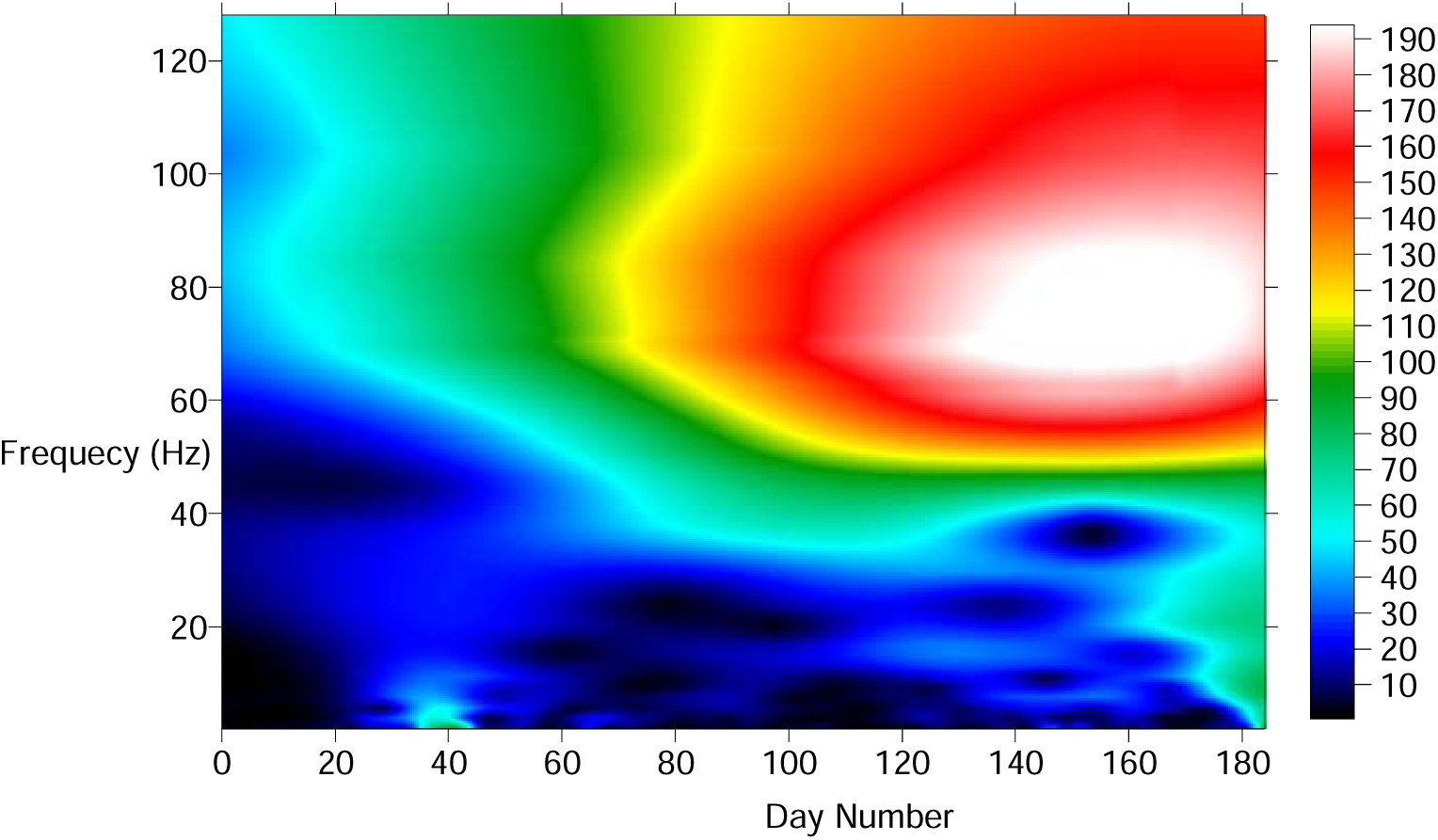
Modulus of the continuous wavelet transform of the daily cases in Algeria

**Figure 3.c.**
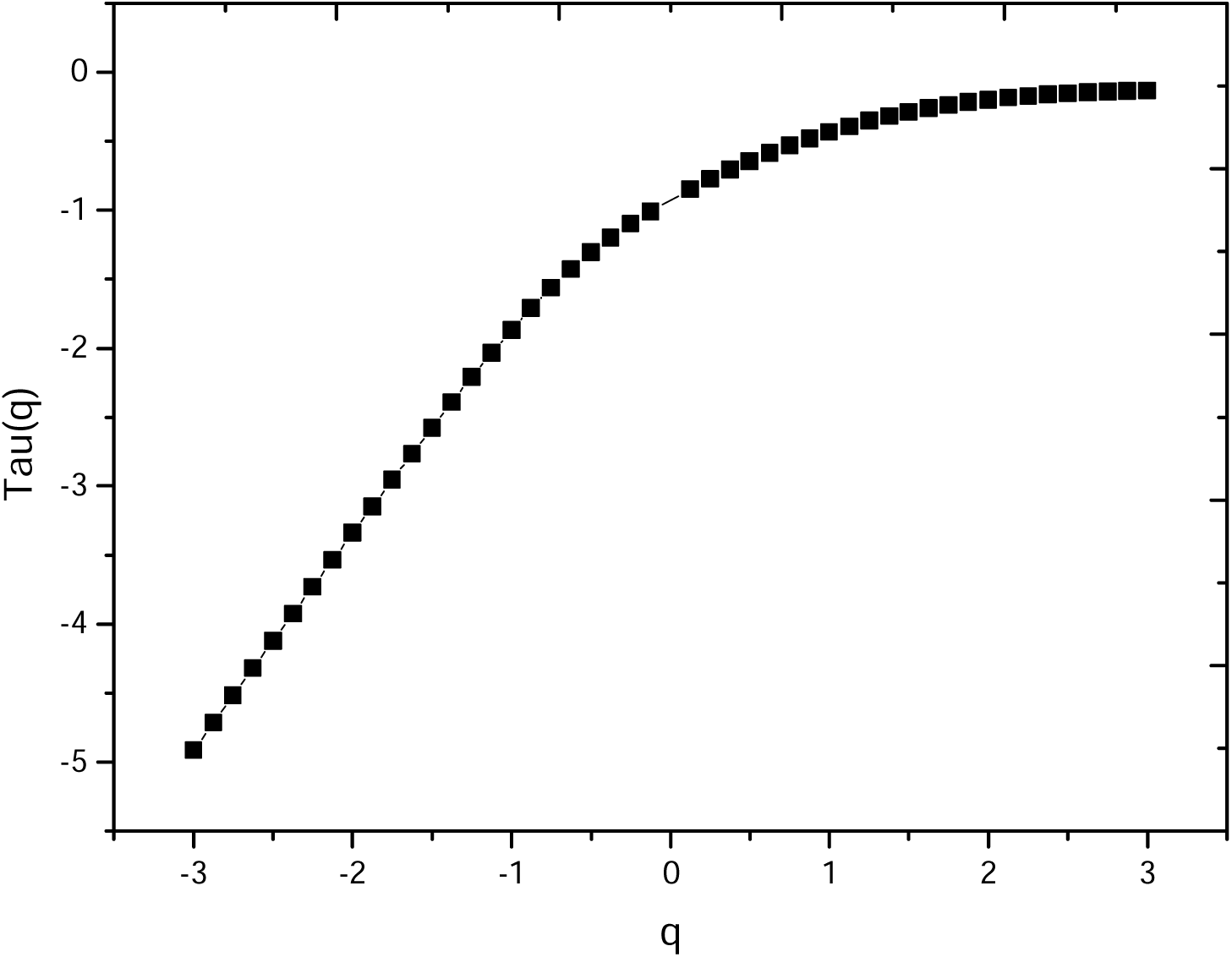
Spectrum of exponents of the daily cases of Algeria

**Figure 1.d.**
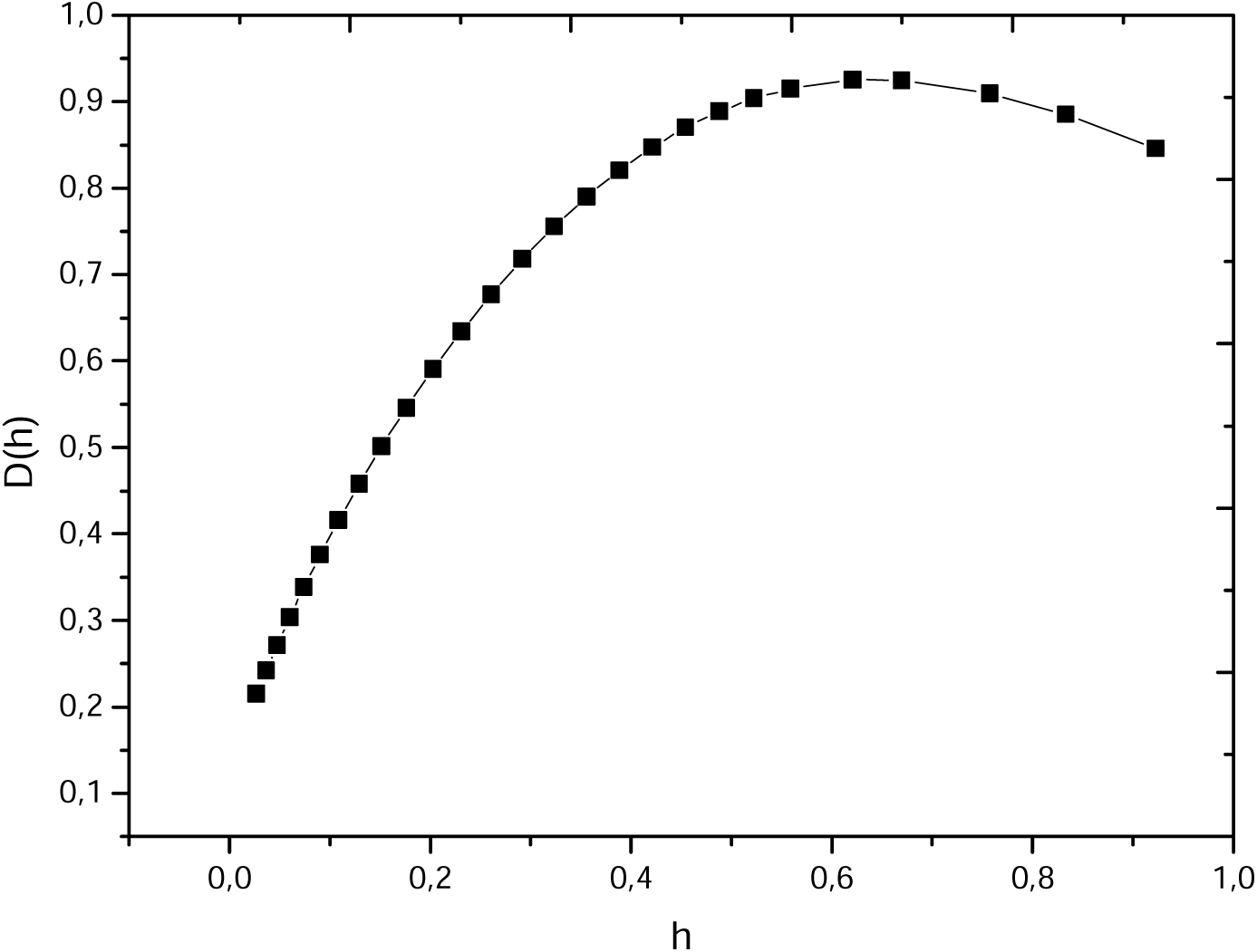
Spectrum of singularities of the daily cases of Algeria

The COVID-19 daily cases in Algeria pandemic fellow a randomized Gaussian model:

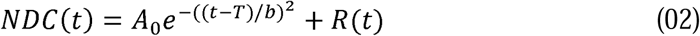

Where *T* = 152, *A*_0_ = 675

*b*: define the lobe opening, for determination of *b* we choose a point *t*_O_ in the curve with *R* (*t*_0_) ≈ 0, For example for the day 125 the daily cases number is 283.

So:

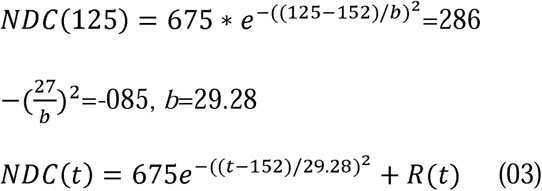

Figure 2.a shows the daily death in Algeria, we can observe that me maximum number of deaths was between the 35 and 50 days from the first observed case, after that there is a stability (between 5-15 cases) in the daily death. Figure 2.b shows that modulus of the continuous wavelet transform in the time-frequency domain.

**Figure 2.a.**
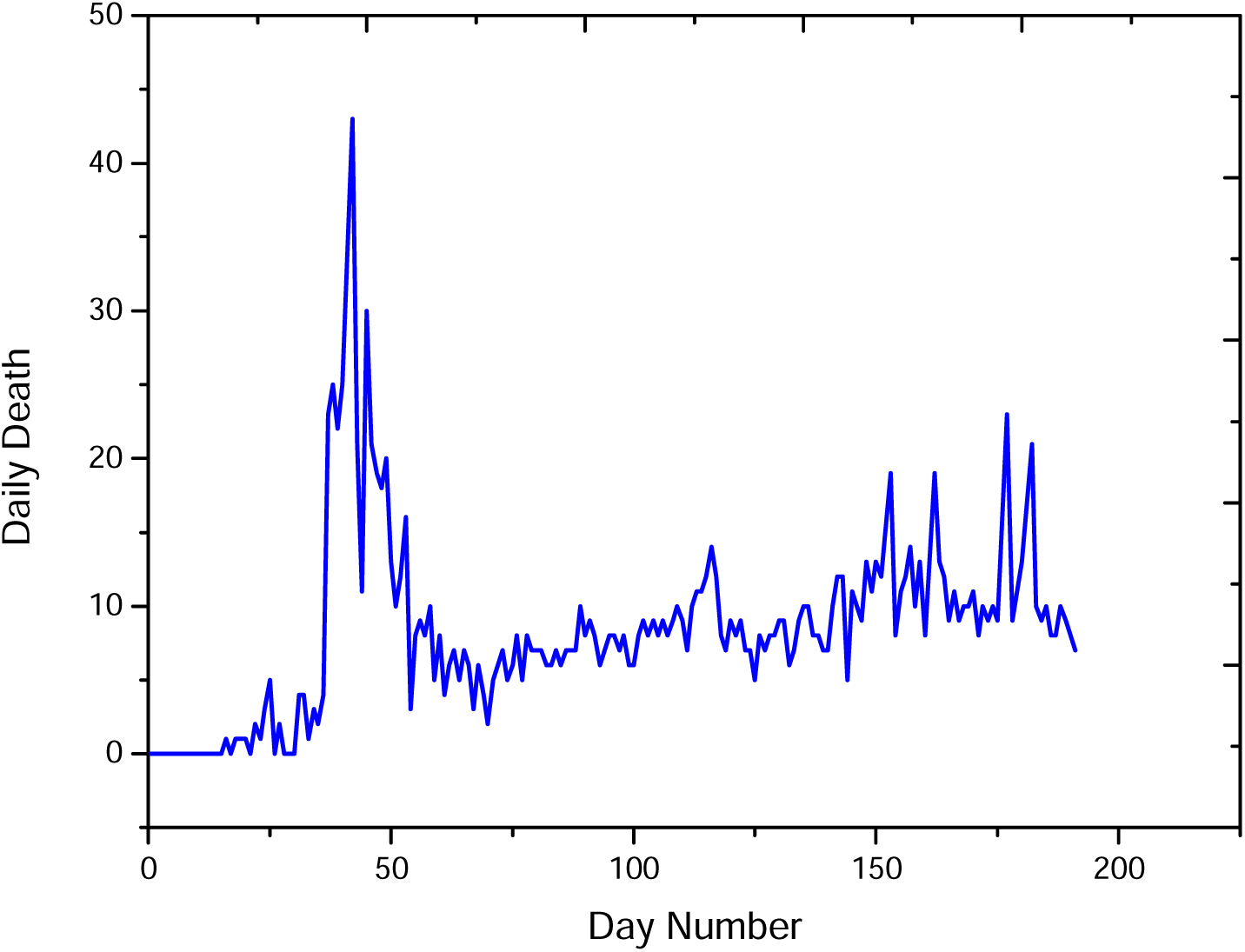
Daily death in Algeria

**Figure 2.b.**
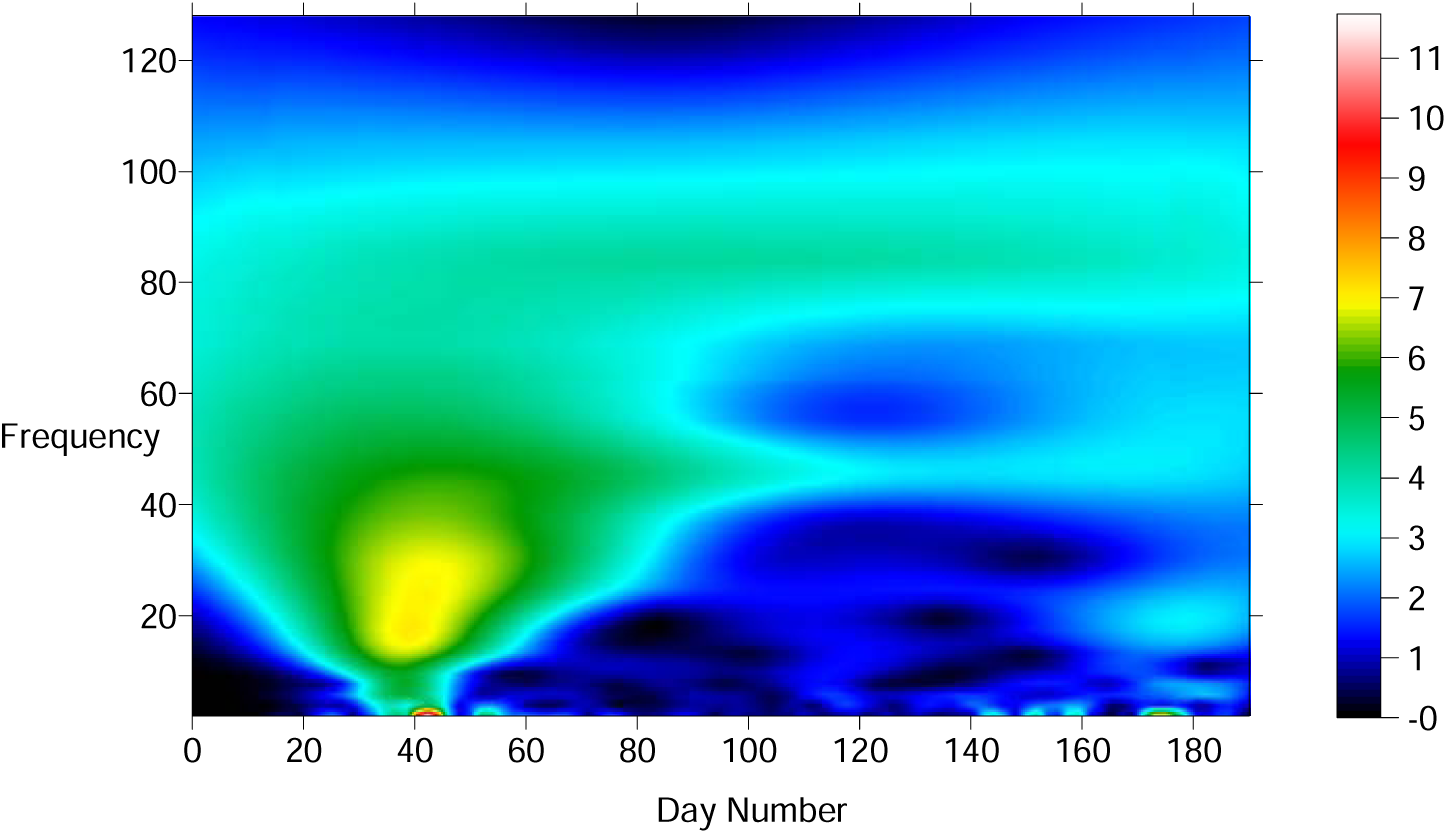
Modulus of the continuous wavelet transform of the daily death in Algeria

Figure 2.c shows the spectrum of exponents, it has a nonlinear behavior demonstrating the multifractal behavior of the daily death cases. Figure 2.d shows the spectrum of singularities, it has a hyperbolic shape with dominant low Holder exponents. This is due the variation in the health sate and immunity of infected patients (chronic illness such as diabetes, arterial hypertension and pneumonic illnesses).

**Figure 2.c.**
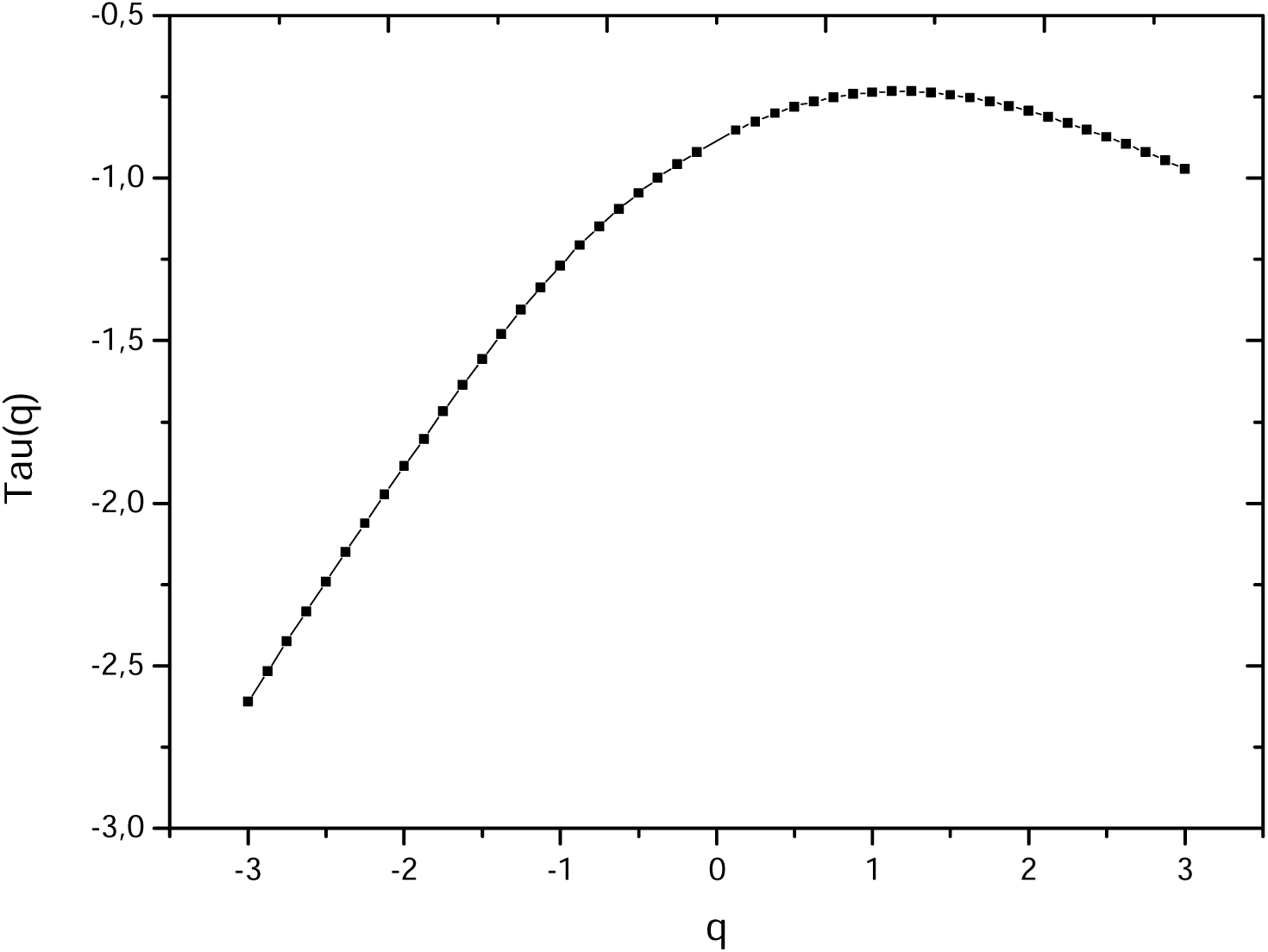
Spectrum of exponents of the daily death of Algeria

**Figure 2.d.**
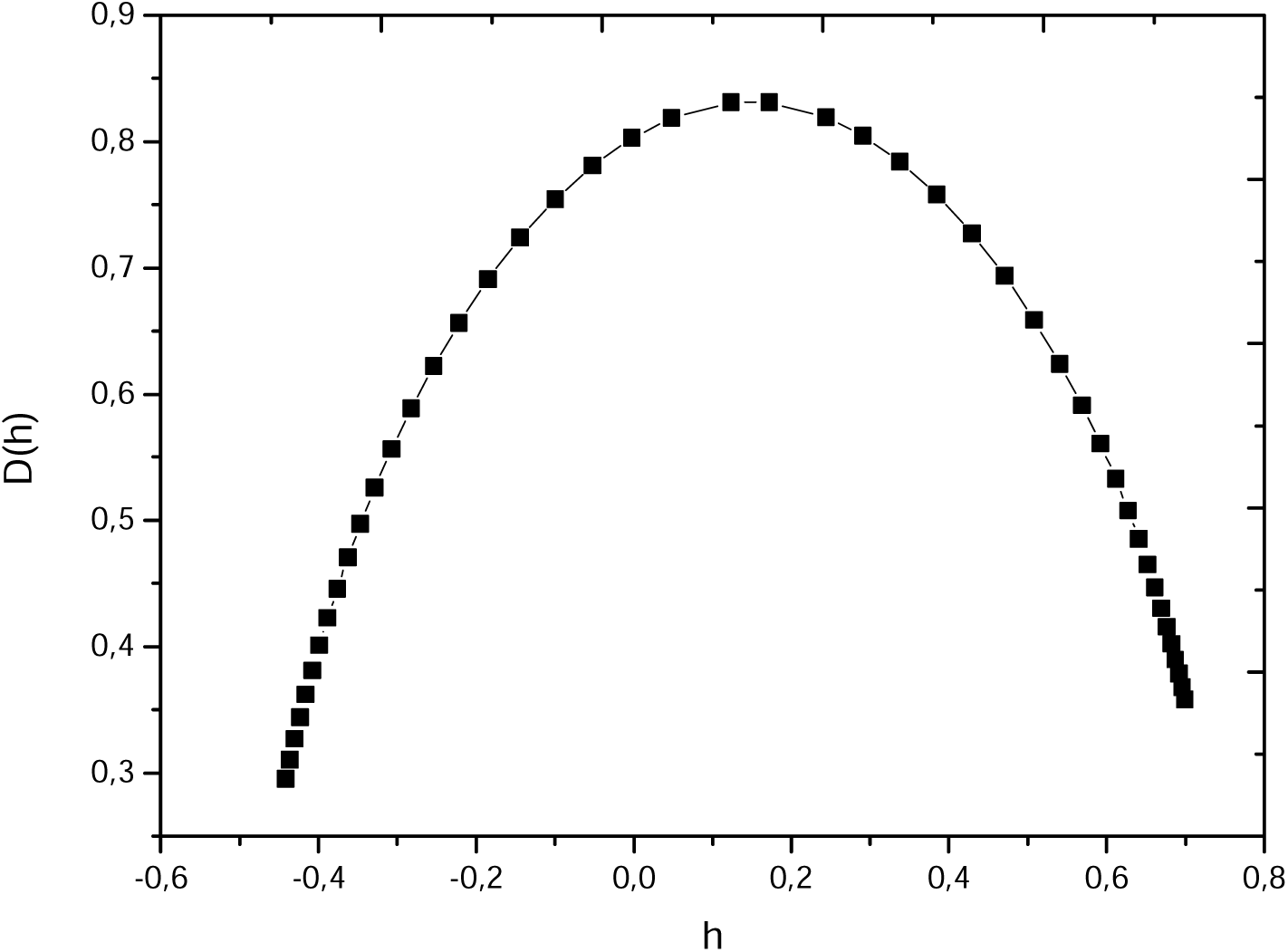
Spectrum of singularities of the daily death in Algeria

### Case of USA

Figure 3.a shows the daily cases in USA, the number of infection is higher that Algeria (this is due to the higher number of population and confinement). Figure 3.b shows the modulus of the continuous wavelet transform in (t-f) domain, while figure 3.c shows the graph of the spectrum of exponents which confirm the multifractal behavior of the daily cases in this country. Figure 3.c shows the spectrum of singularities versus the Holder exponents, we observe that the Holder exponents are very low (<0.53), which confirm the presence of the high frequency components only in the daily cases signal. The presence of this character in the SARS-CoV COVID-19 pandemic spread in USA is due to the non-respect of confinement in this country. The first observed case was in 20^th^ of January 2020, we observe also that the SARS-CoV2 coronavirus has the maximum aggressiveness between June and September 2020. The propagation looks like will become more stable in the future. Figure 4.a shows the graph of the daily death in USA, we can observe that the maximum death number was during the period of April-June 2020 this may be due to the version the SARS-CoV2 coronavirus that was very dangerous and fatal in this period, since the virus is far from stability and the equilibrium and it undergoes a lot of mutation process (Raoult, 2020; Mandal, 2020; Ouadfeul, 2020).

**Figure 3.a.**
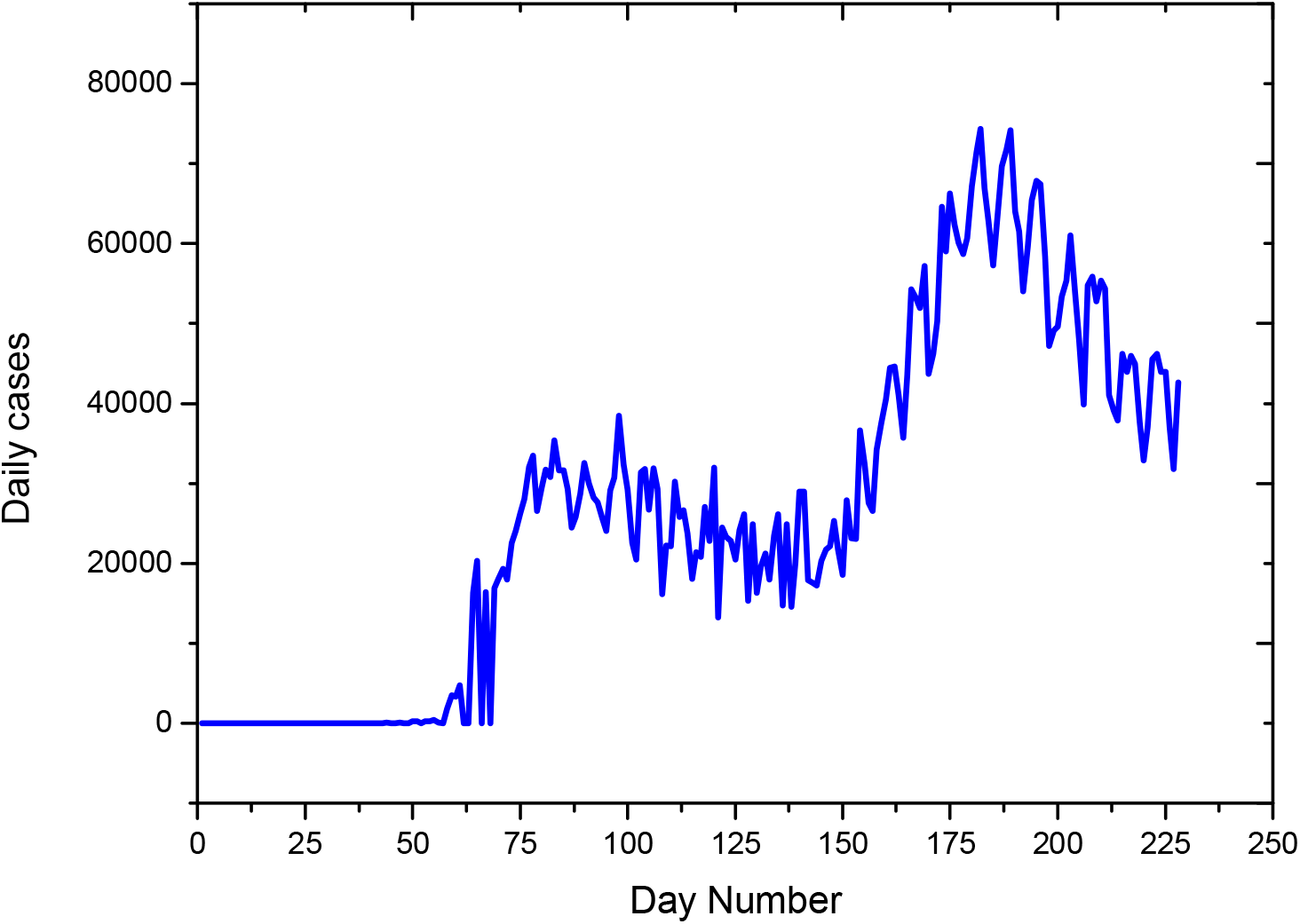
Daily cases in USA

**Figure 3.b.**
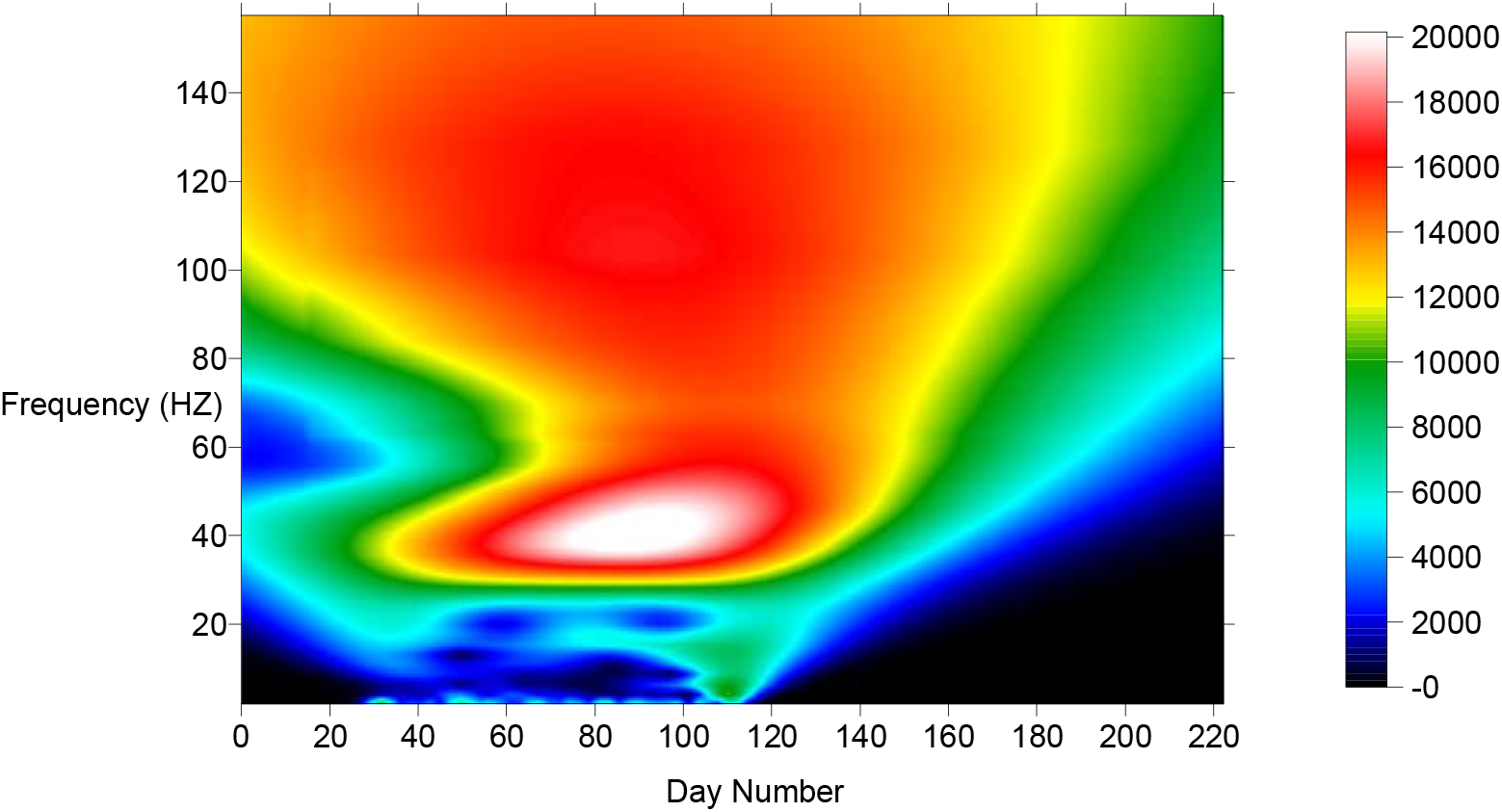
Modulus of the continuous wavelet transform of the daily cases in USA

**Figure 3.c.**
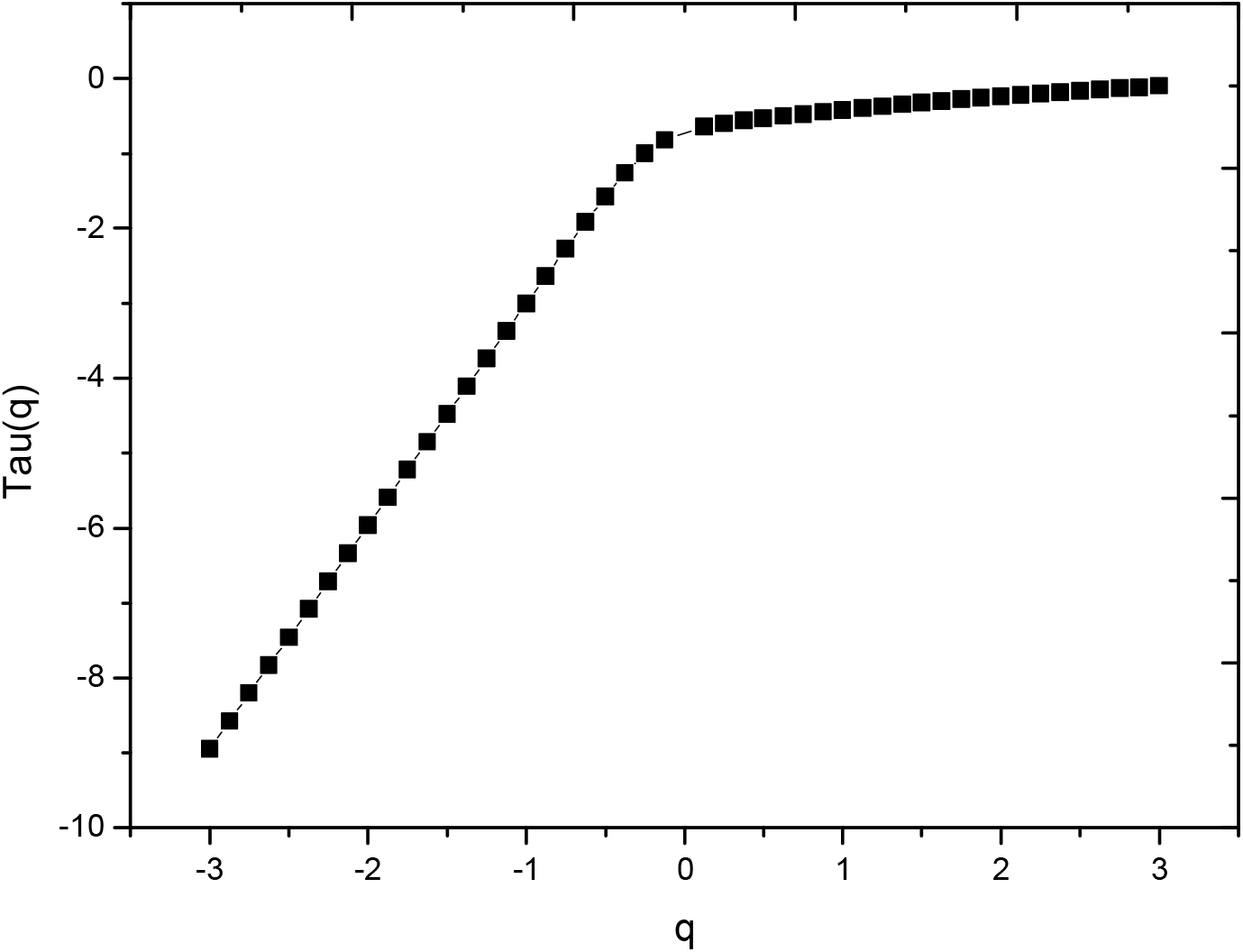
Spectrum of exponents of the daily cases in USA

**Figure 3.d.**
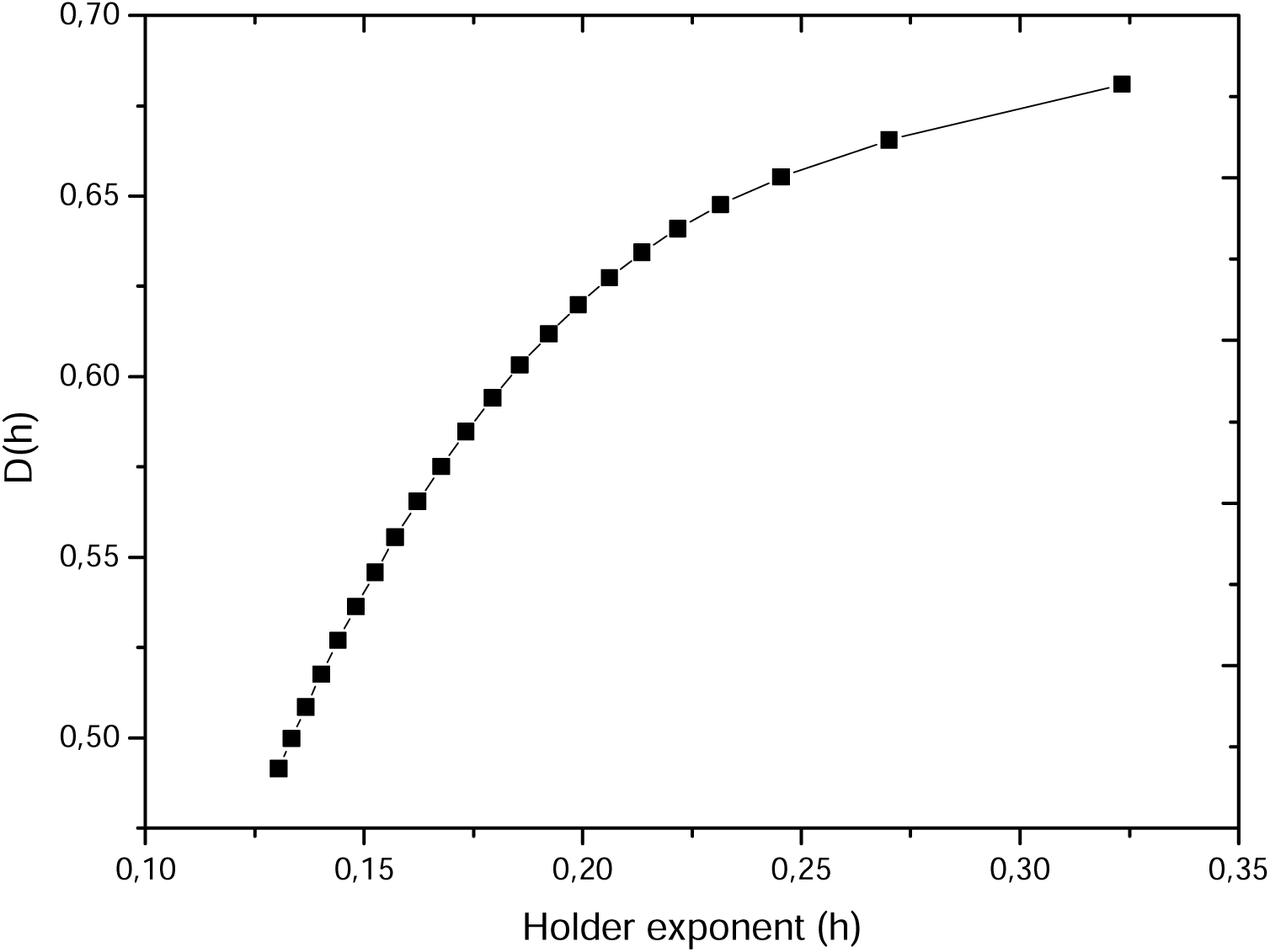
Spectrum of singularities of the daily cases in USA

**Figure 4.a.**
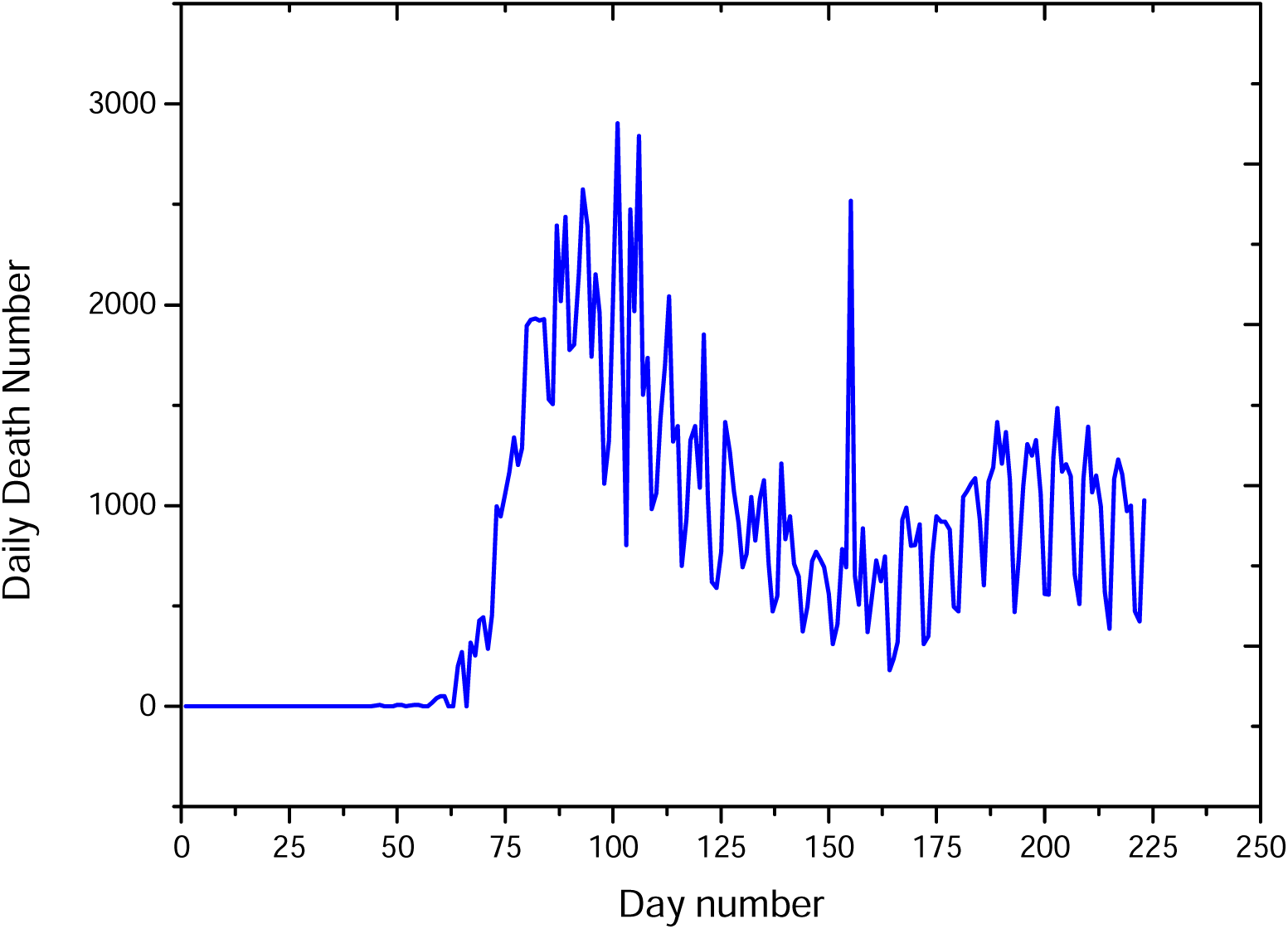
Daily death in USA

**Figure 4.b.**
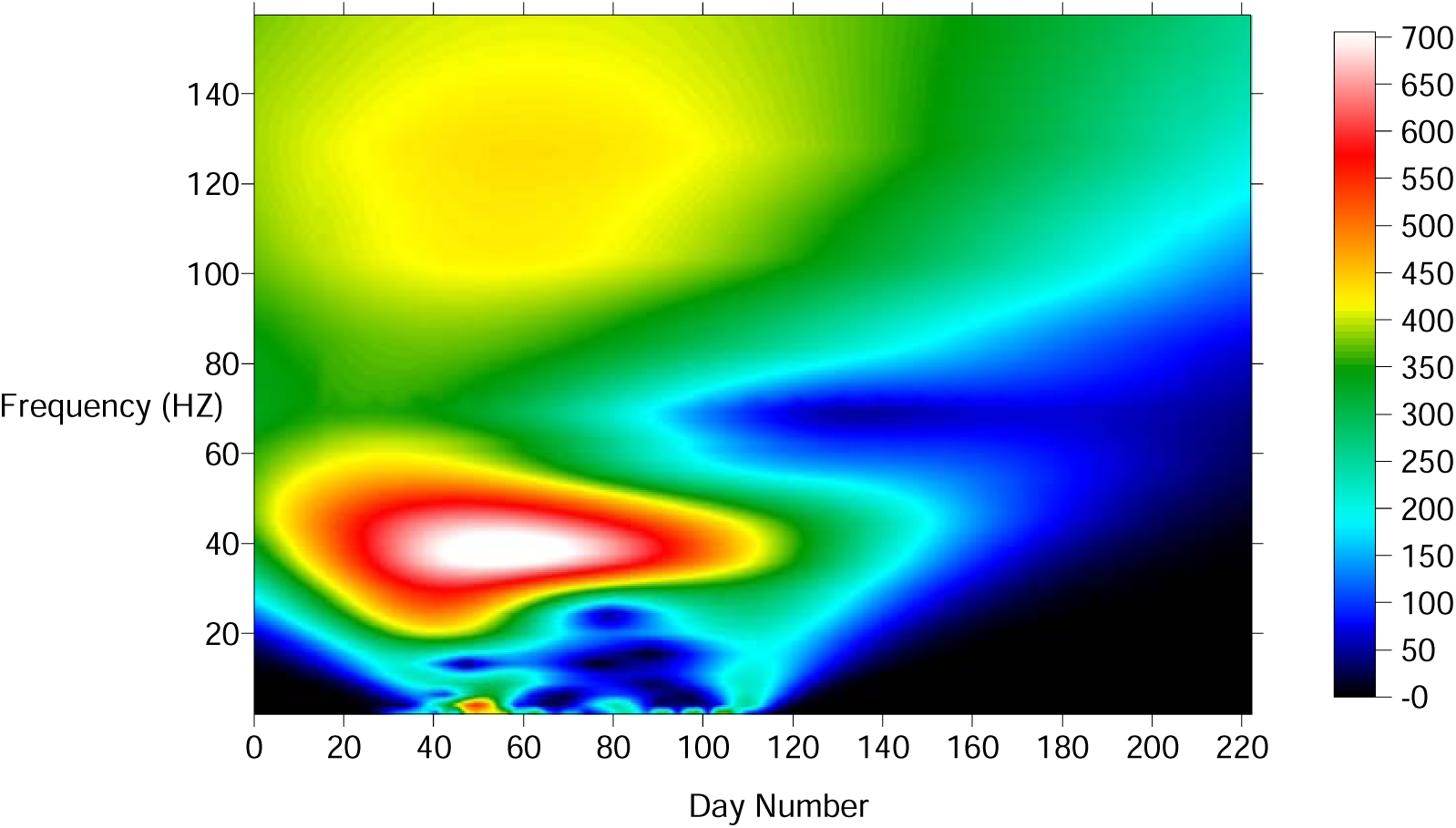
Modulus of the continuous wavelet transform of the daily death in USA

**Figure 4.c.**
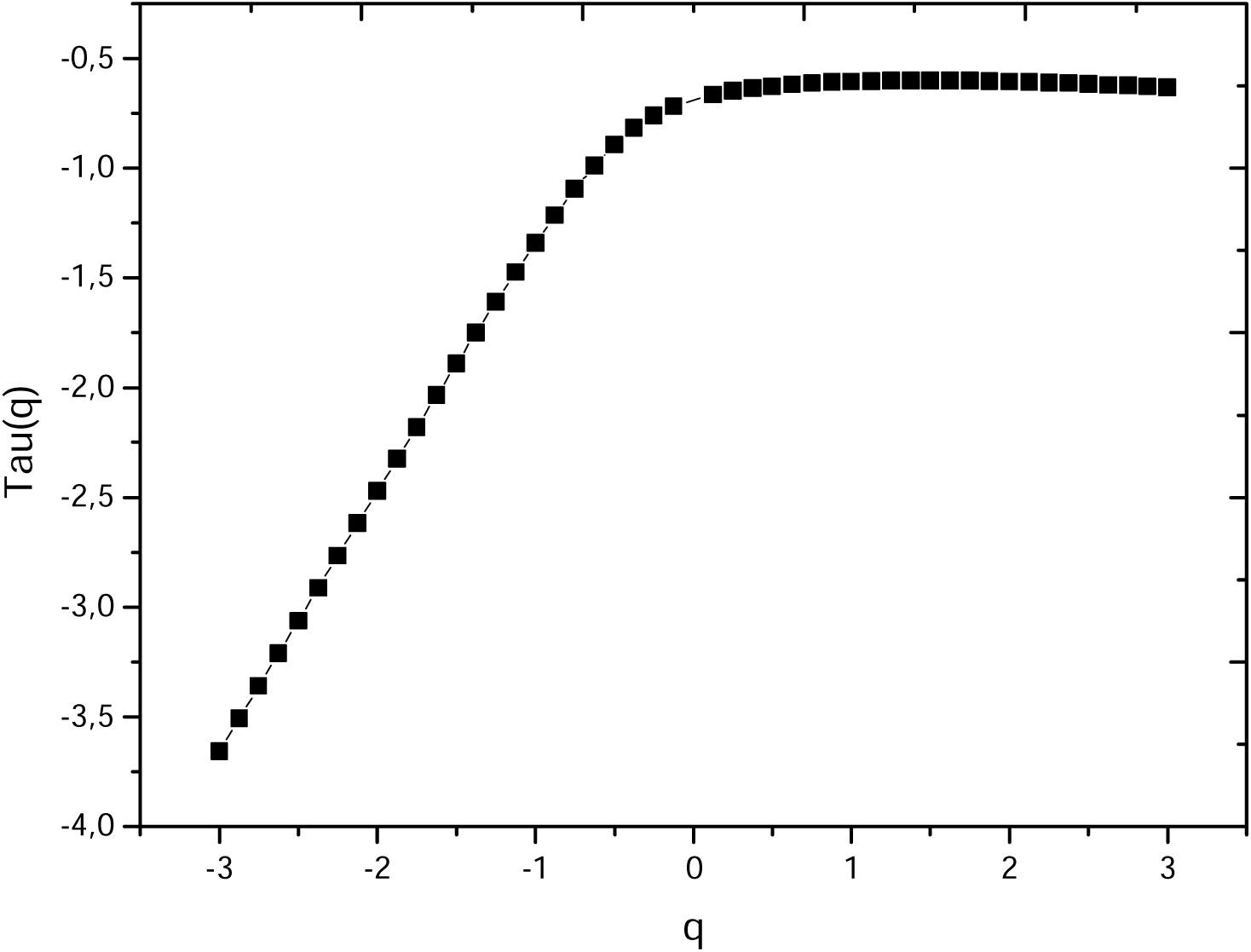
Spectrum of exponents of the daily death in USA

**Figure 4.d.**
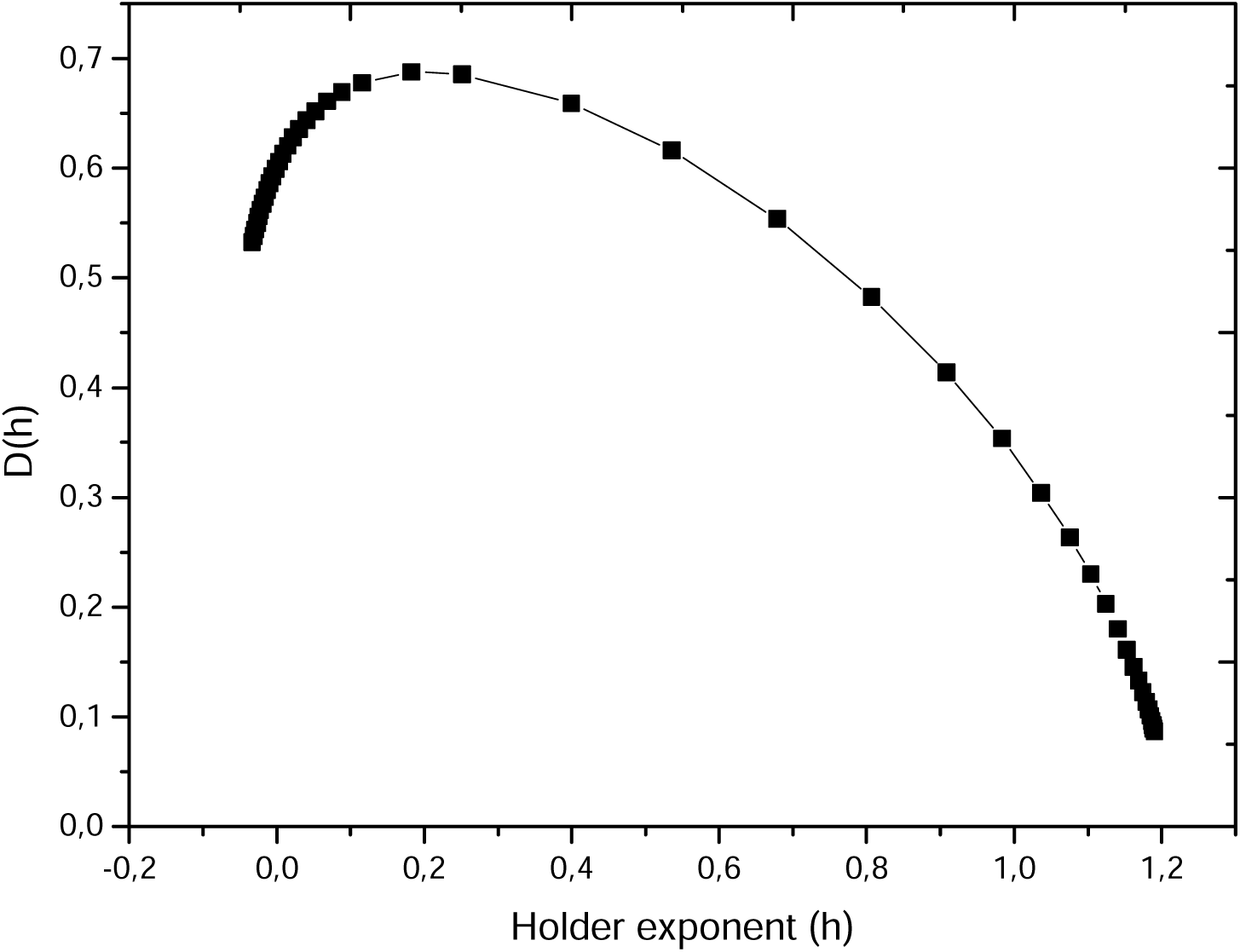
Spectrum if singularities of the daily death in USA

### Case of Russia

Figure 5.a shows the graph of daily cases in Russia, it is in decrease. The first observed case was in the 31^th^ of January 2020, the maximum number of cases is between May-September 2020. Figure 5.b shows the modulus of the continuous wavelet transform, the analyzing wavelet is the complex Morlet, while figure 5.c shows the spectrum of exponents which exhibits a nonlinear behavior demonstrating the multifractal behavior of COVID-19 daily cases signal. Figure 5.d shows the graph of the spectrum of singularities versus the Holder exponents, we observe the dominance of the range of Holder exponents between [0.4-1] which confirm the dominance of medium to low frequencies components in the daily cases signal. This is due to the progressive decrease in the pandemic daily decrease.

**Figure 5.a.**
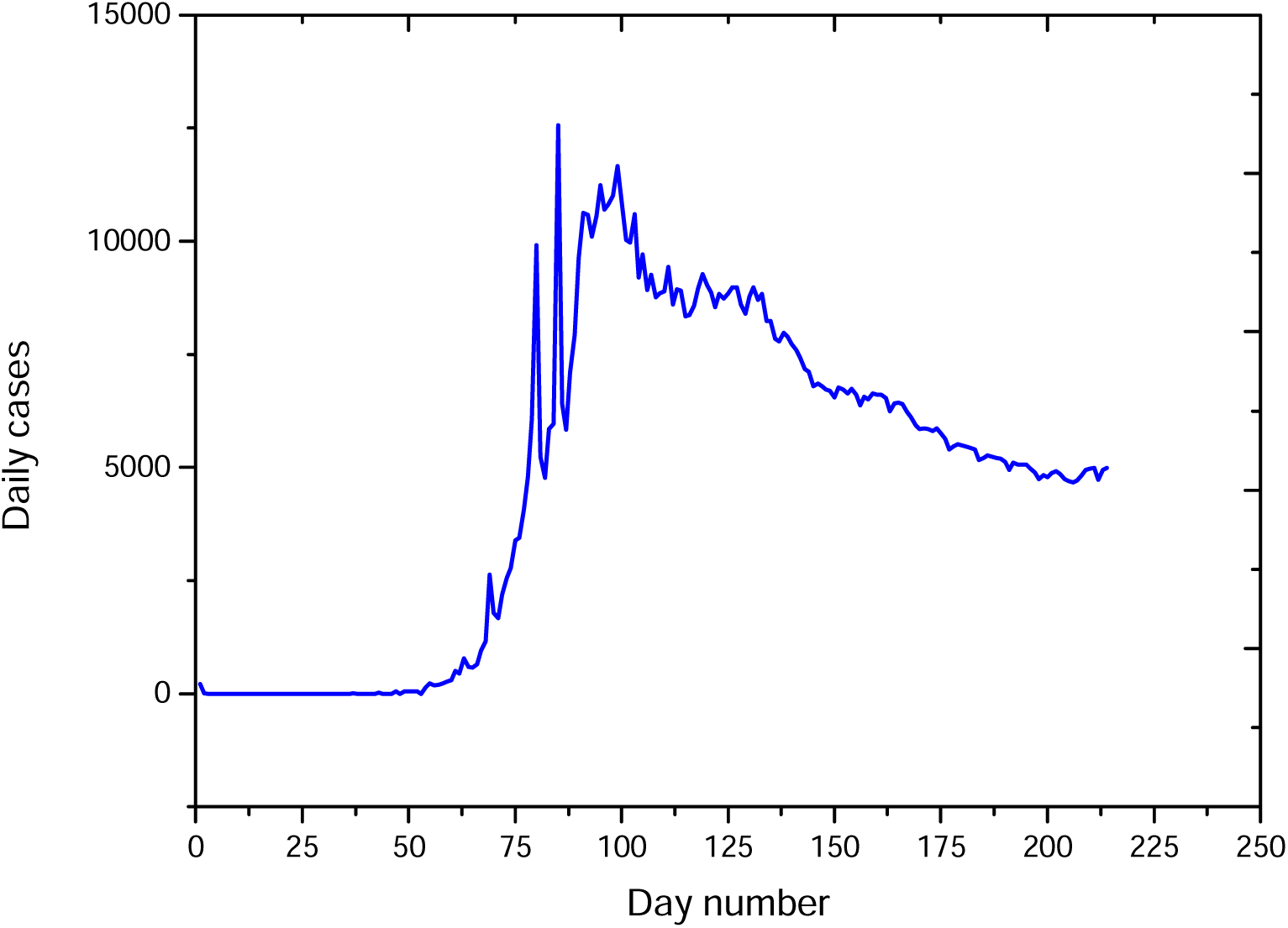
Daily cases in Russia

**Figure 5.b.**
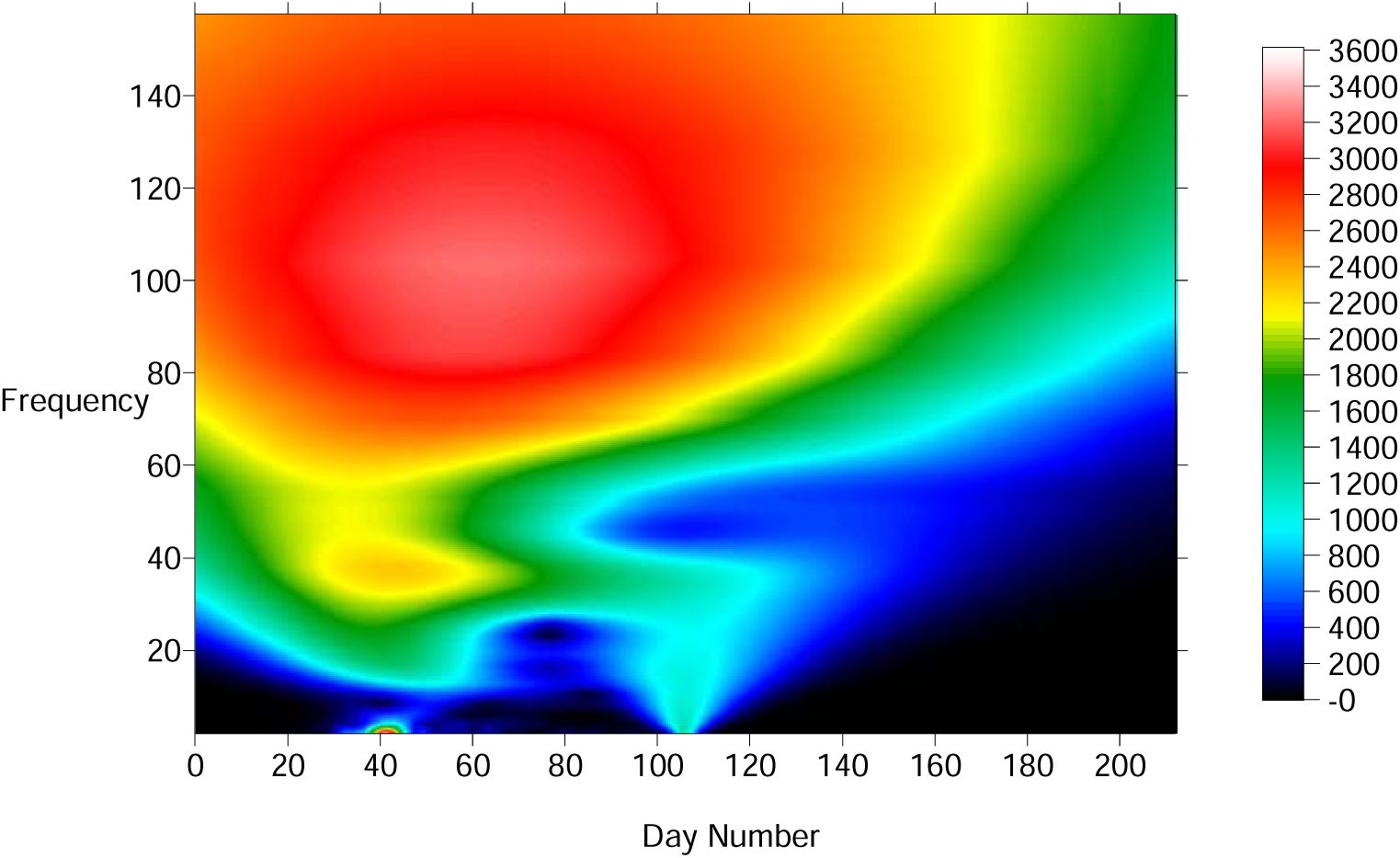
Modulus of the continuous wavelet of the daily cases in Russia.

**Figure 5.c.**
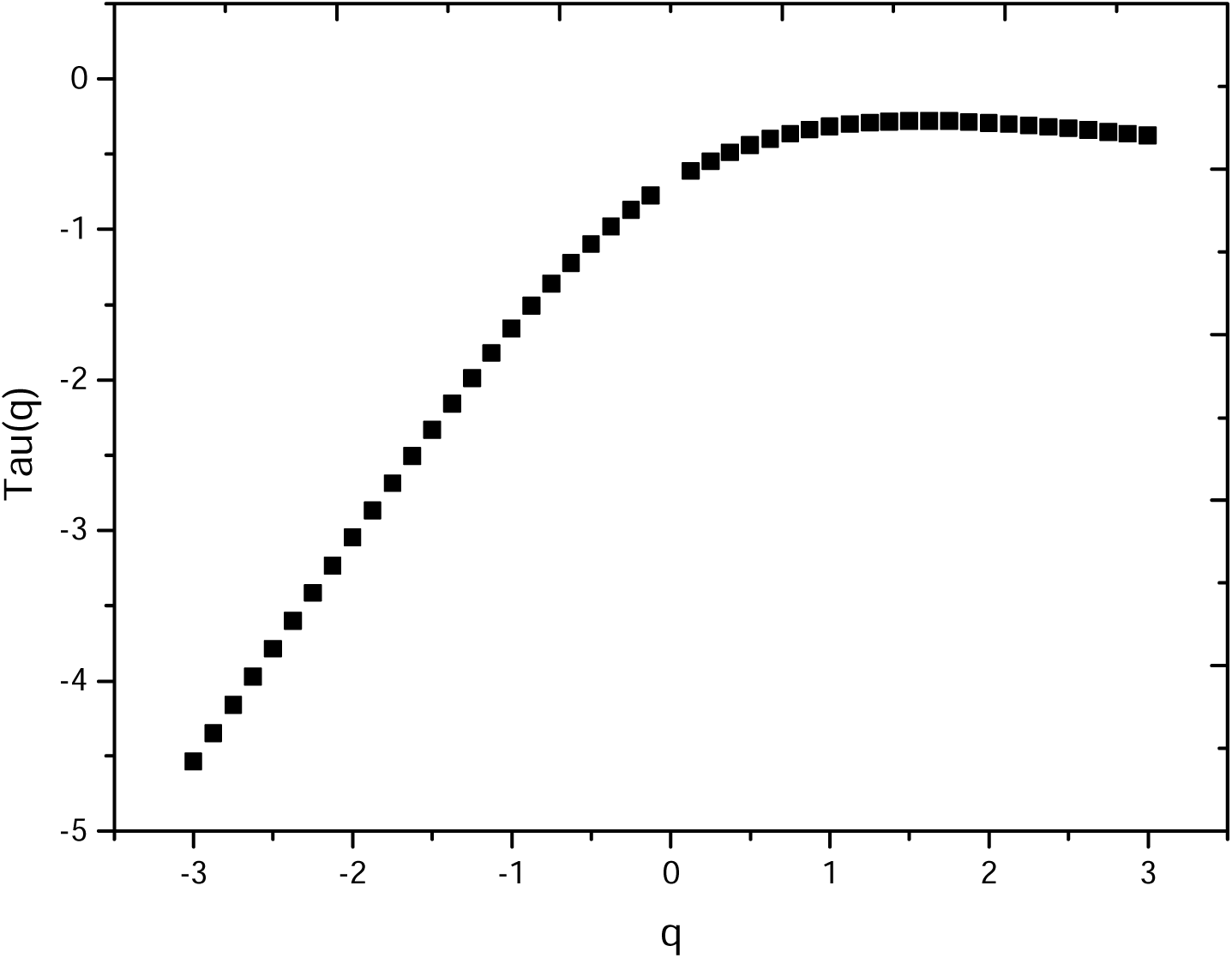
Spectrum of exponents of the daily cases in Russia.

**Figure 5.d.**
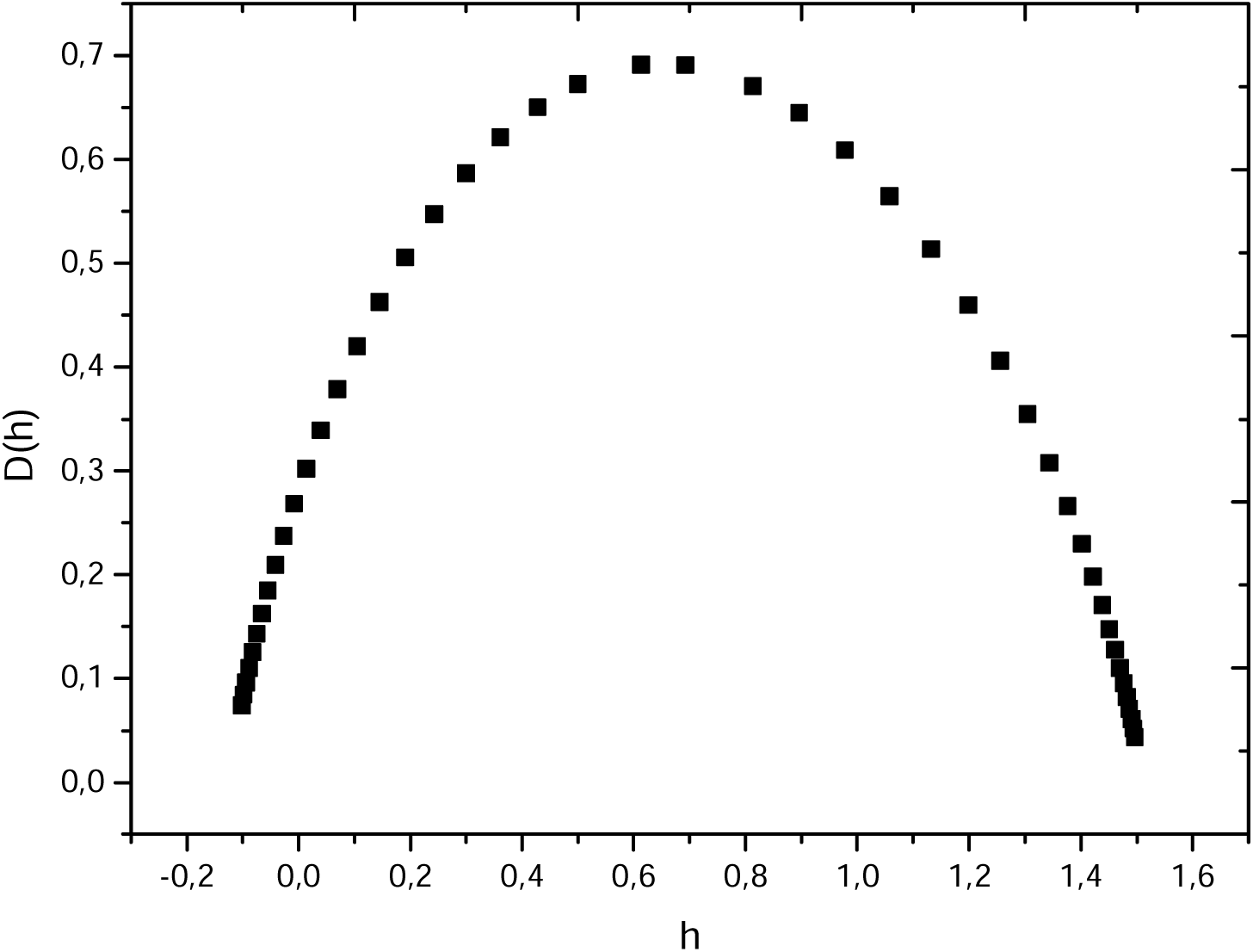
Spectrum of singularities of the daily cases in Russia.

Figure 6.a shows the graph of the daily death in Russia, it looks like a randomized modified Takagi model, the number of daily death is in decrease. Figure 6.b shows the modulus of the continuous wavelet transform while figure 6.c shows the spectrum of exponents which a nonlinear behavior confirming the multifractal behavior of the daily death in Russia.

Figure 6.d shows the graph the spectrum of singularities, the dominance of the low Holder exponents is clear. The high variation in the daily death number due to the variation in the health state of the infected people (presence/absence of chronic illnesses).

**Figure 6.a.**
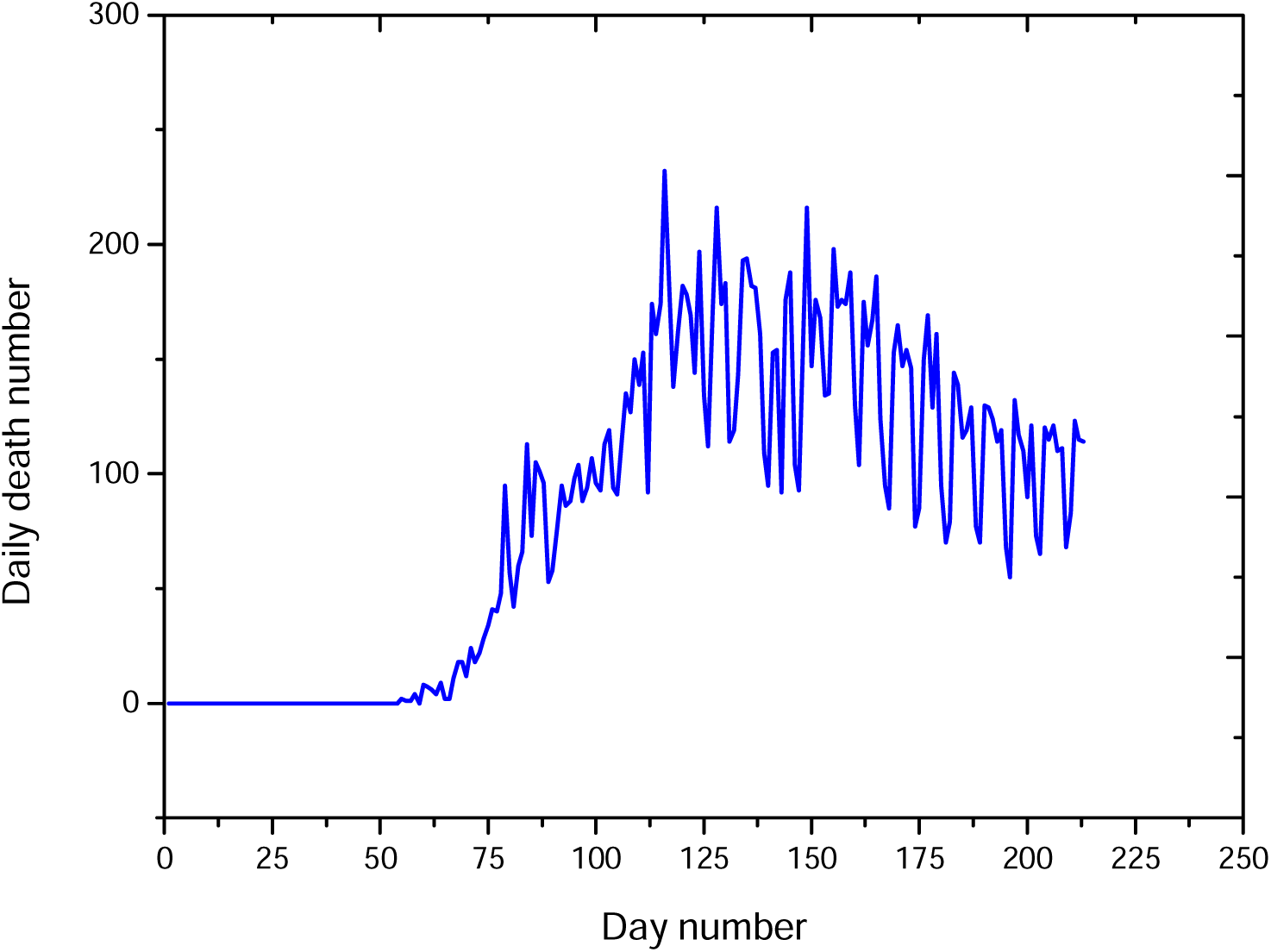
Daily death in Russia

**Figure 6.b.**
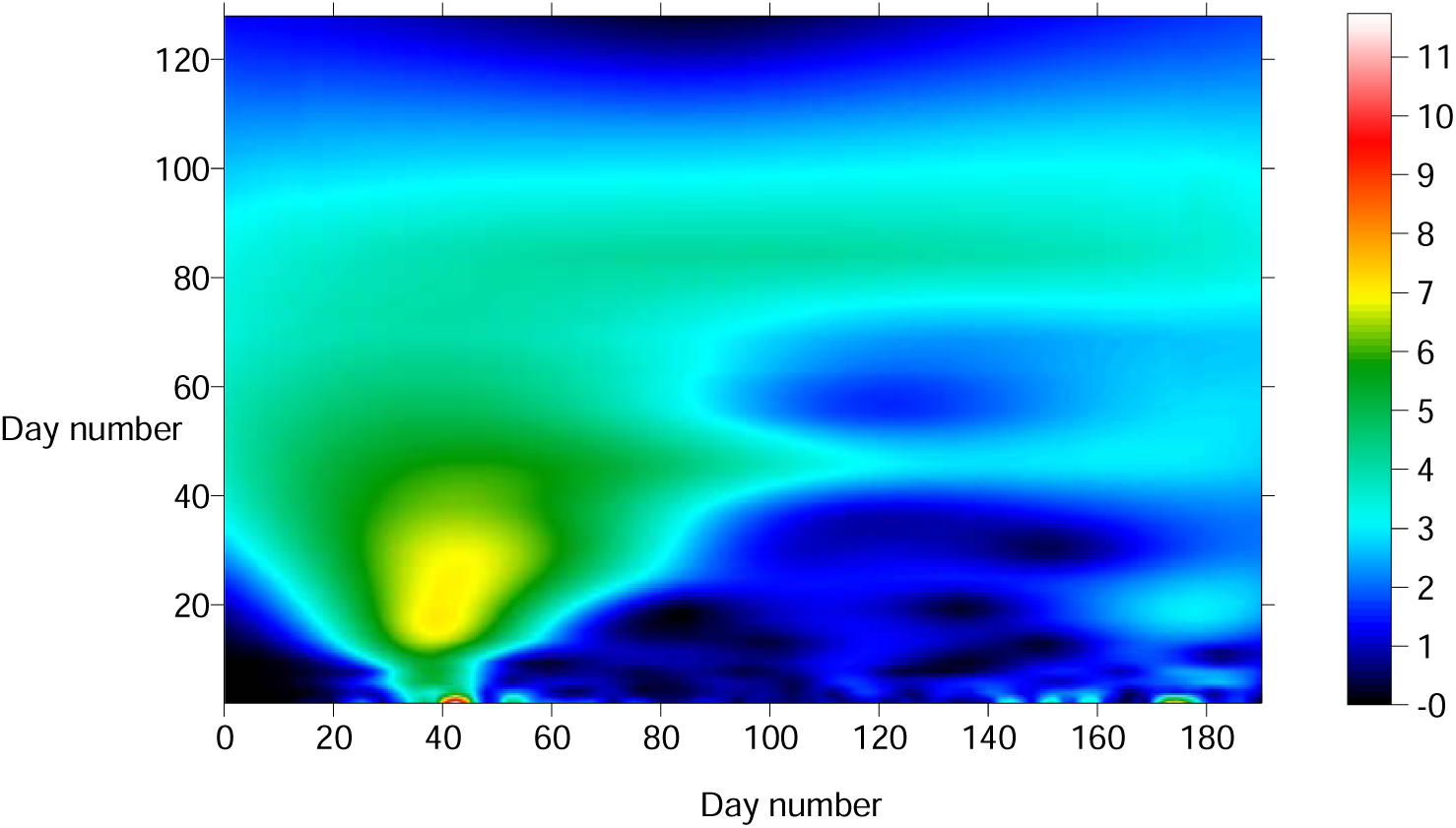
Modulus of the continuous wavelet transform in the (time-frequency) domain of daily death in Russia

**Figure 6.c.**
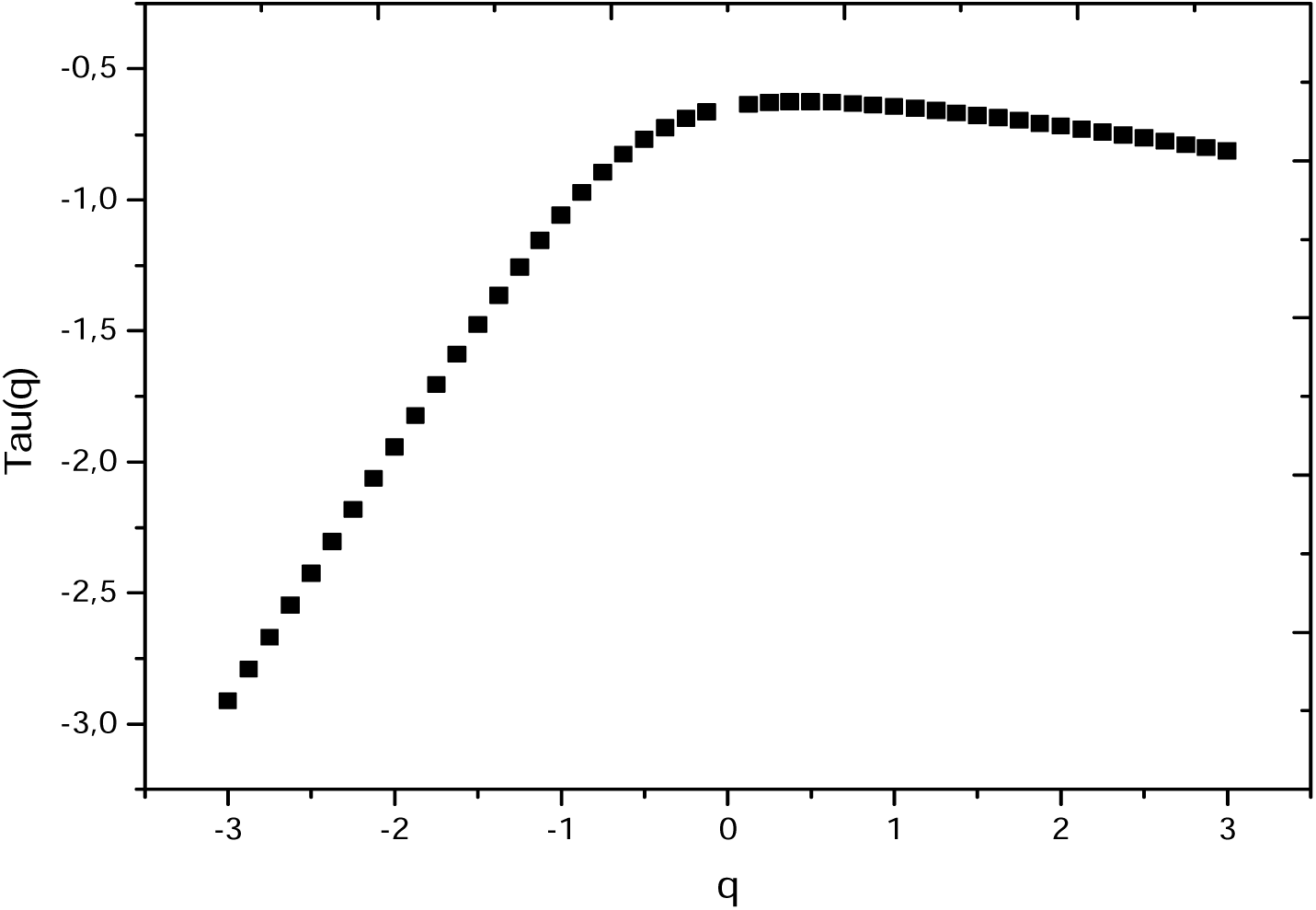
Spectrum of exponents of daily death in Russia

**Figure 6.d.**
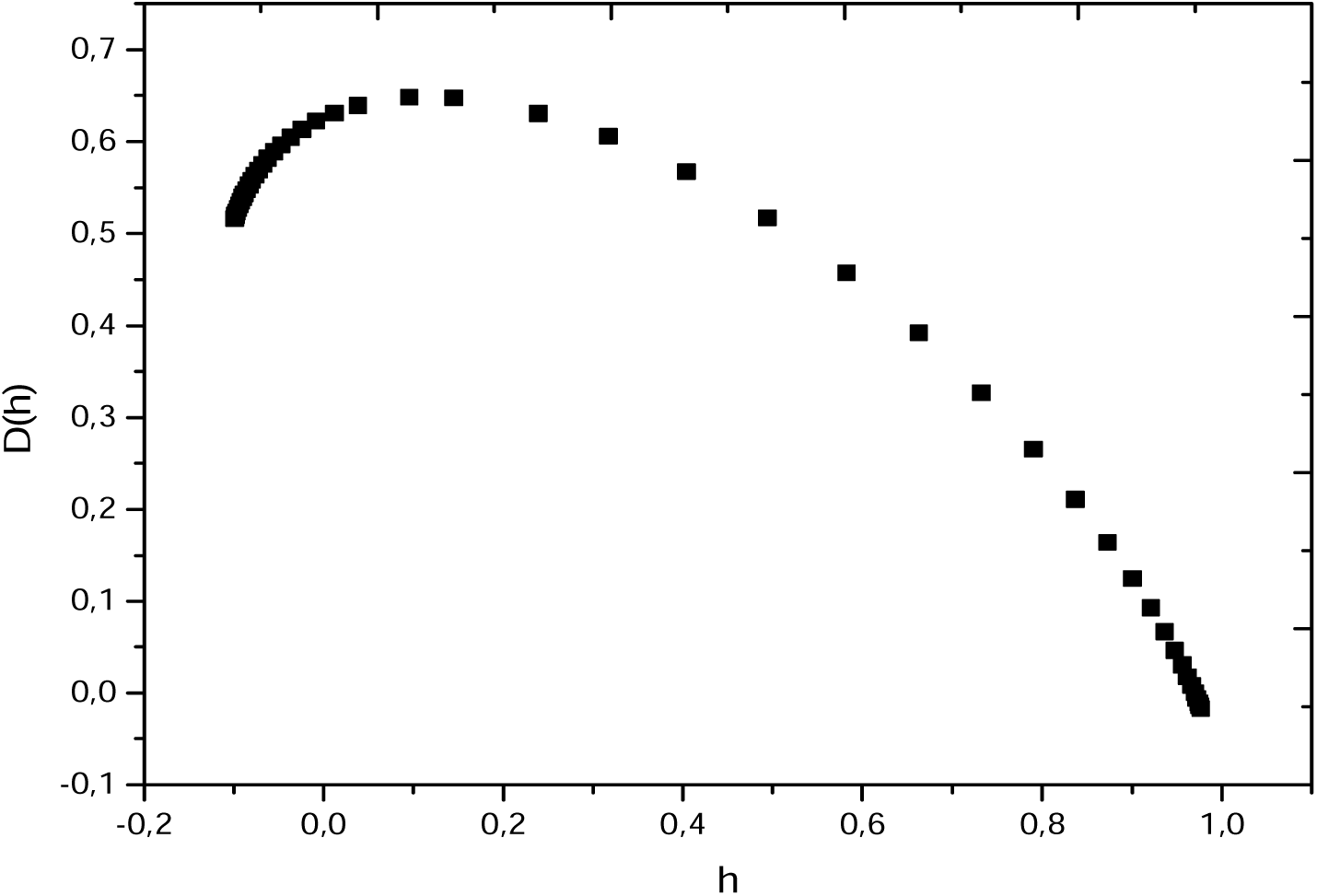
Spectrum of singularities of the daily death in Russia.

### Case of Italy

Figure 7.a shows the COVID-19 daily cases in Italy, the first observed case was in the 29^th^ if January 2020, the maxima of pandemic was between February-May 2020, after that the situation become stable until the beginning of the month of August where a second wave of the pandemic seems to be in the front. Figure 7.b shows the modulus of the continuous wavelet transform, while figure 7.c shows the spectrum of exponents versus *q* which is demonstrating the multifractal behavior of SARS-CoV-2 COVID-19 epidemic in Italy. Figure 7.d shows the spectrum of singularities, we observe the dominance of the high Holder exponents, which means the absence of the high frequency components in this signal. The origin of absence of the high frequency components is the progressive increase7decrease of the daily cases. Figure 8.a shows the daily death of COVID-19 pandemic in Italy, the maximum death number was between Mars and May 2020, after the number of death was very low. This due to the version of SARS-CoV2 coronavirus which is under the mutation process (Raoult, 2020).

**Figure 7.a.**
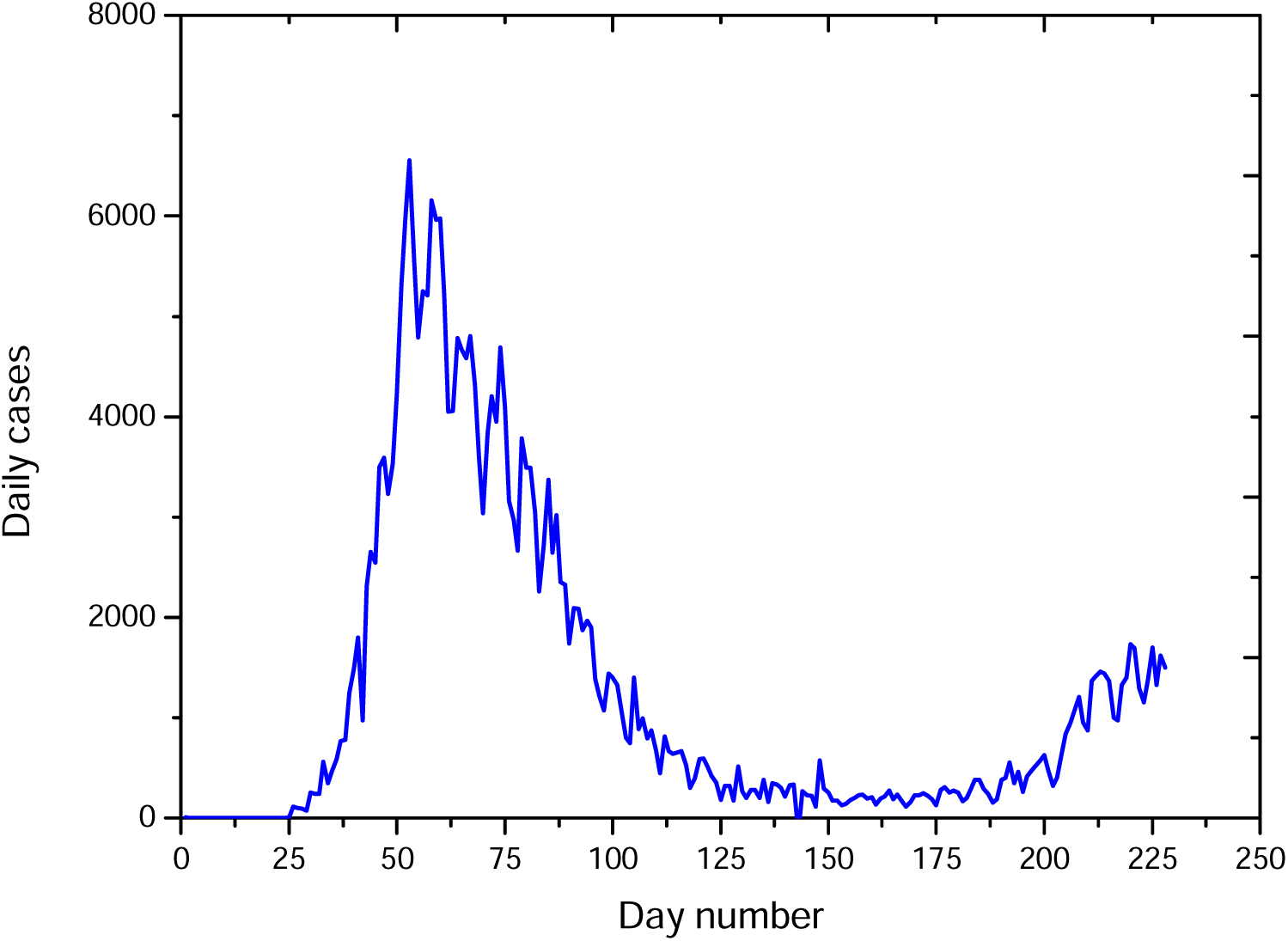
Graph of the daily cases in Italy

**Figure 7.b.**
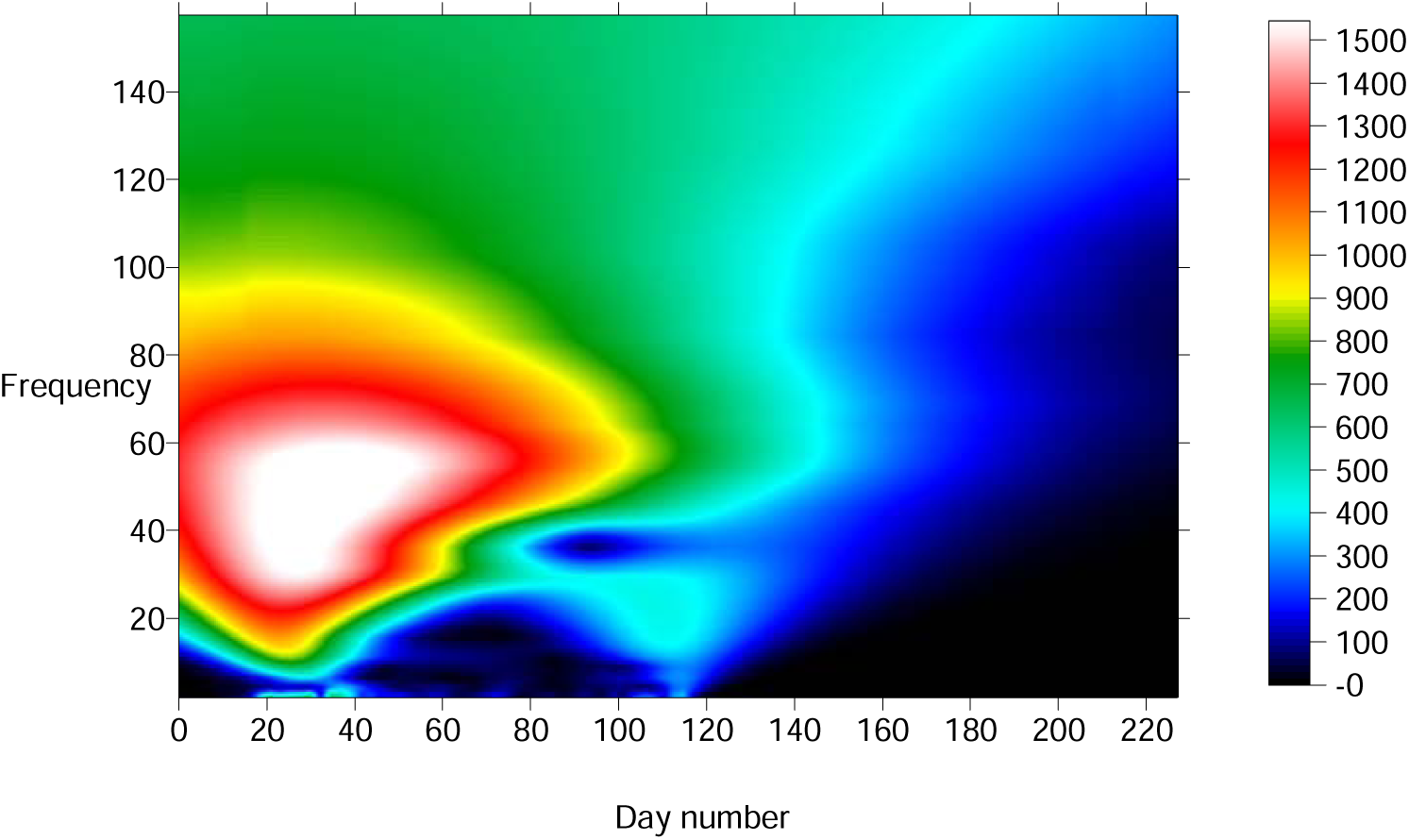
Modulus of the continuous wavelet transform of daily cases in Italy

**Figure 7.d.**
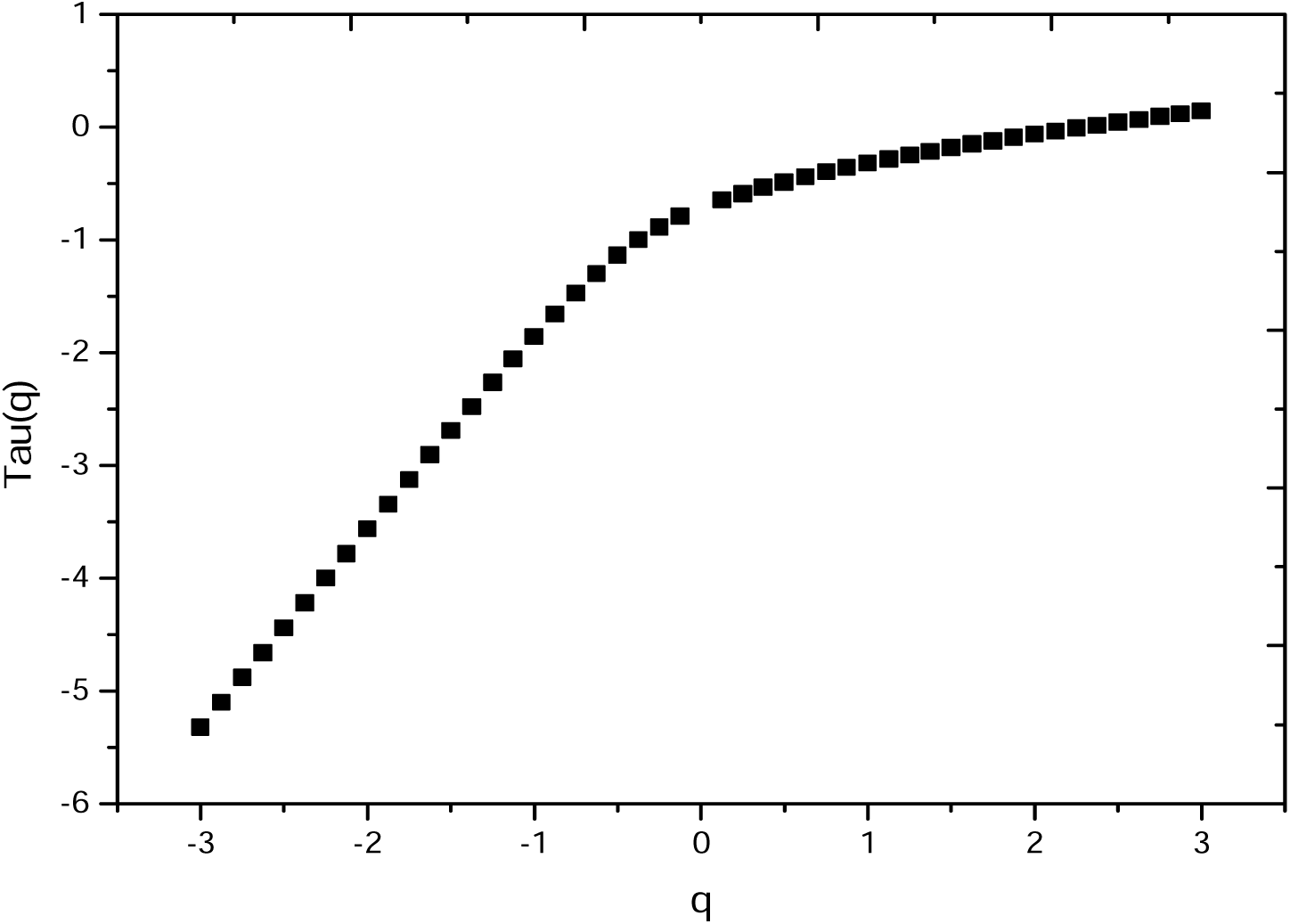
Spectrum of exponents of the daily cases in Italy

**Figure 7.d.**
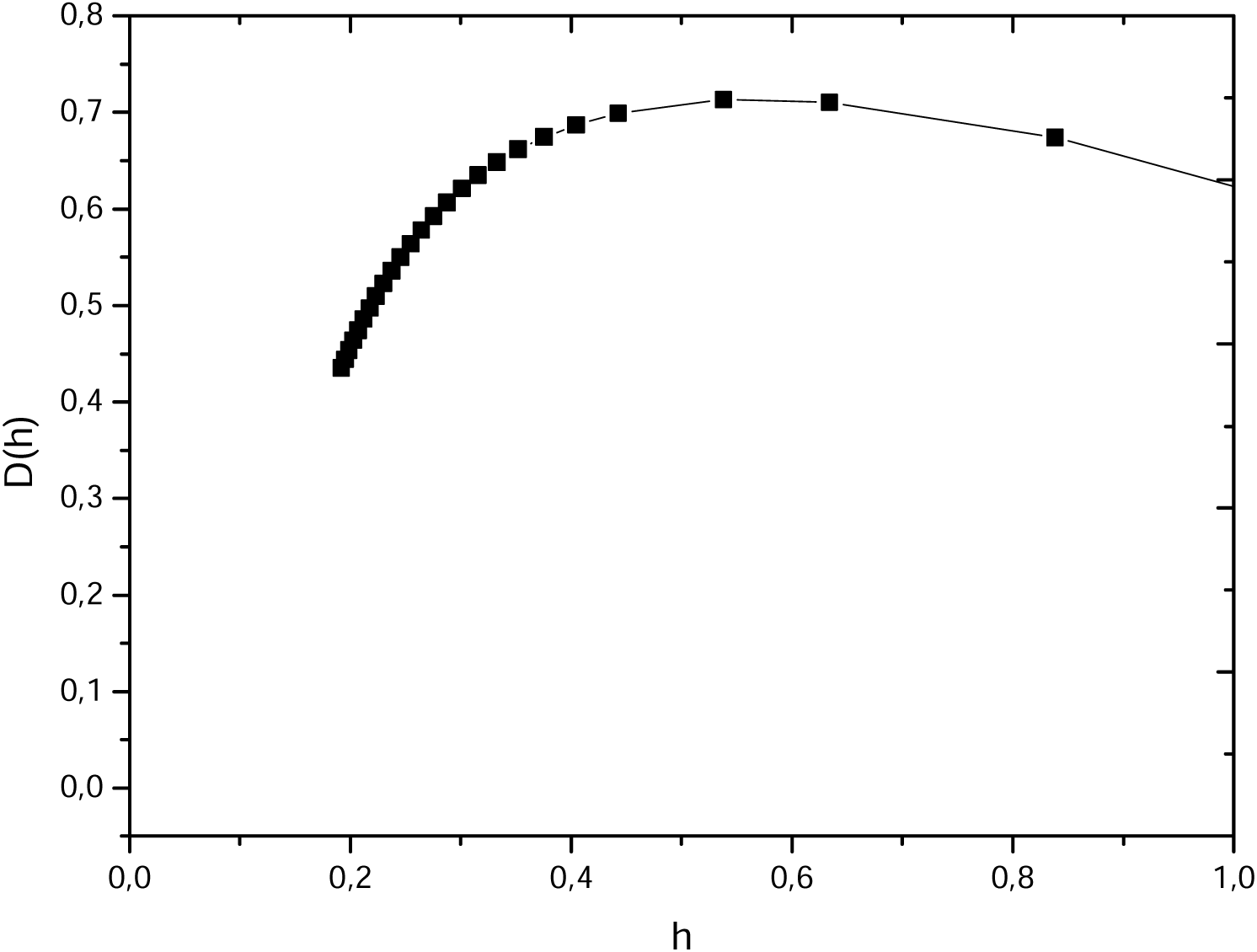
Spectrum of singularities of the daily cases in Italy

Figure 8.b shows the modulus of the continuous wavelet transform in the (time-frequency) domain, while figures 8.c and 8.d show the graph of the spectrum of exponents and the spectrum of singularities, the multifractal behavior of the daily death cases is clearly demonstrated in these spectra. The low roughness coefficients character is dominant in this signal; the source of this characteristic is the variation in the health sate of infected patients.

**Figure 8.a.**
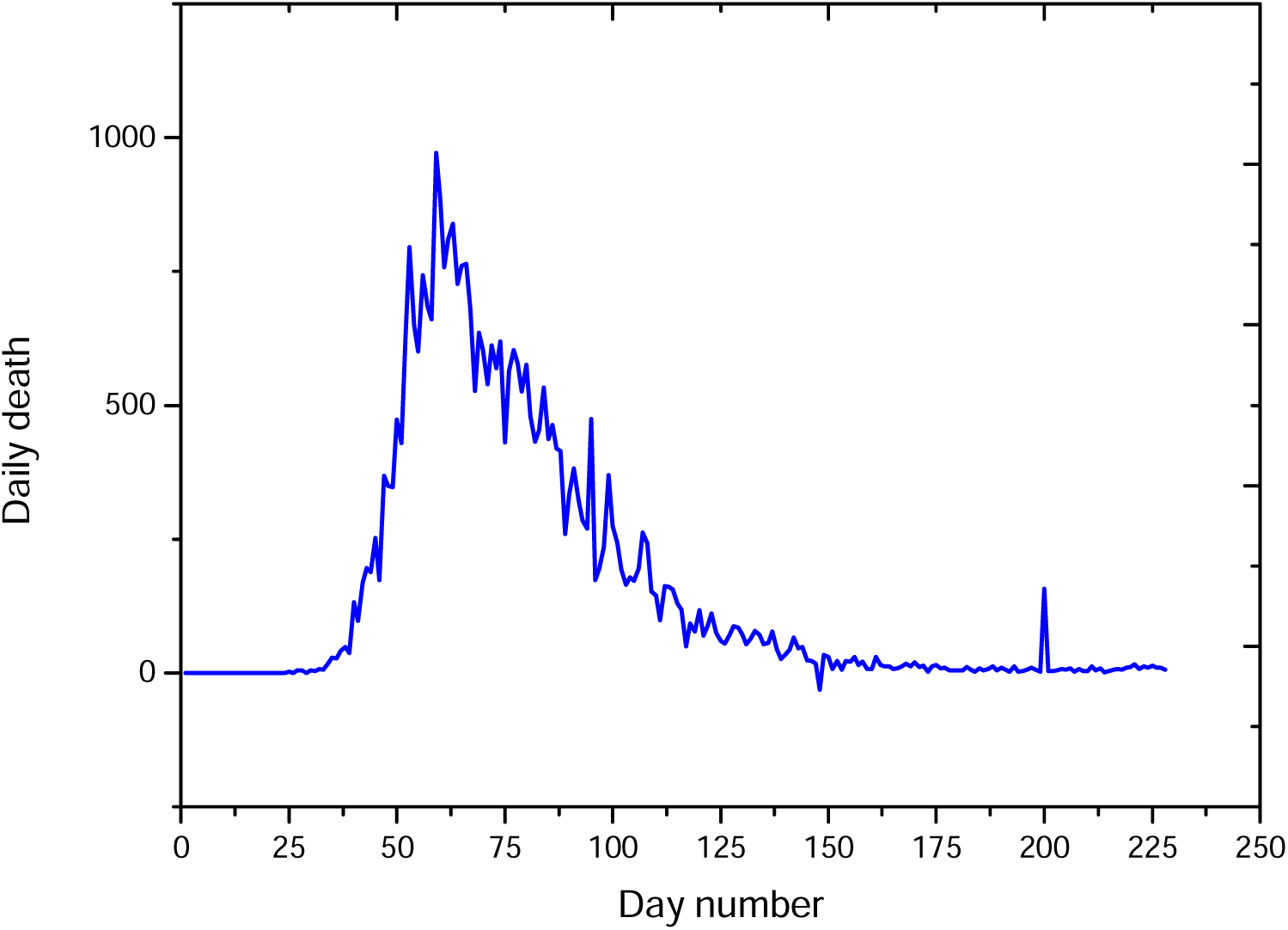
Graph of the daily death in Italy

**Figure 8.b.**
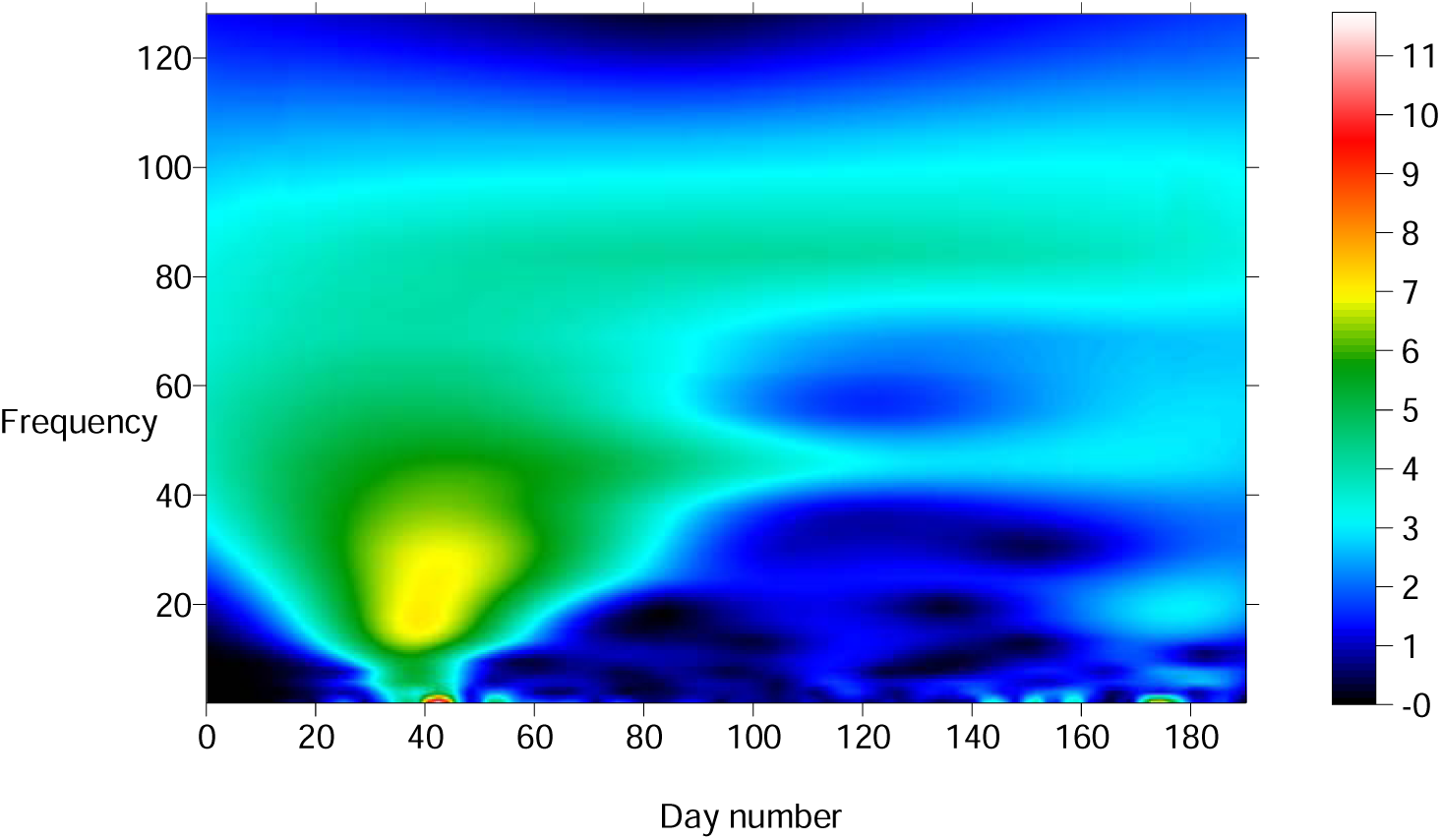
Modulus of the continuous wavelet transform of the daily death in Italy

**Figure 8.c.**
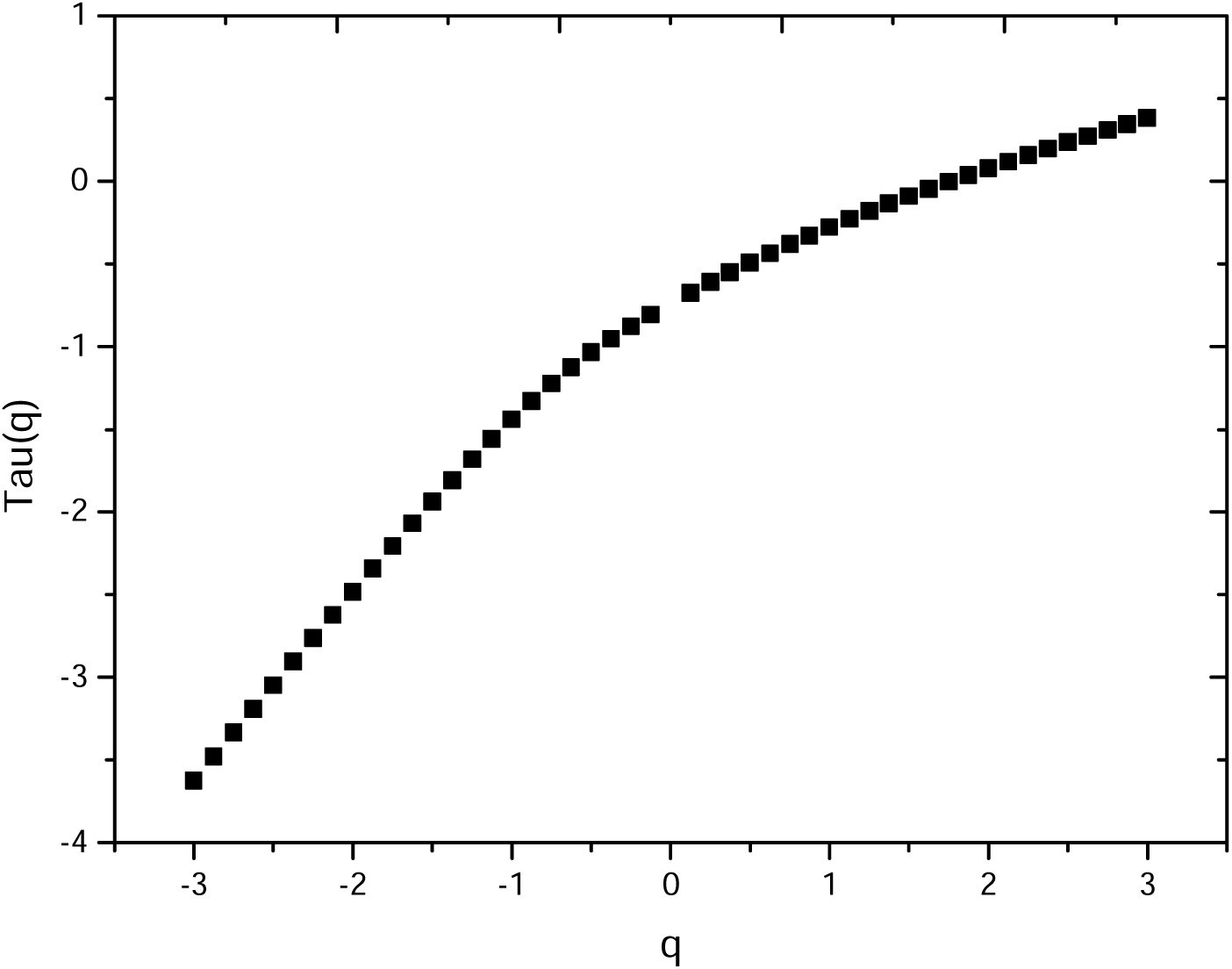
Spectrum of exponents of the daily death in Italy

**Figure 8.d.**
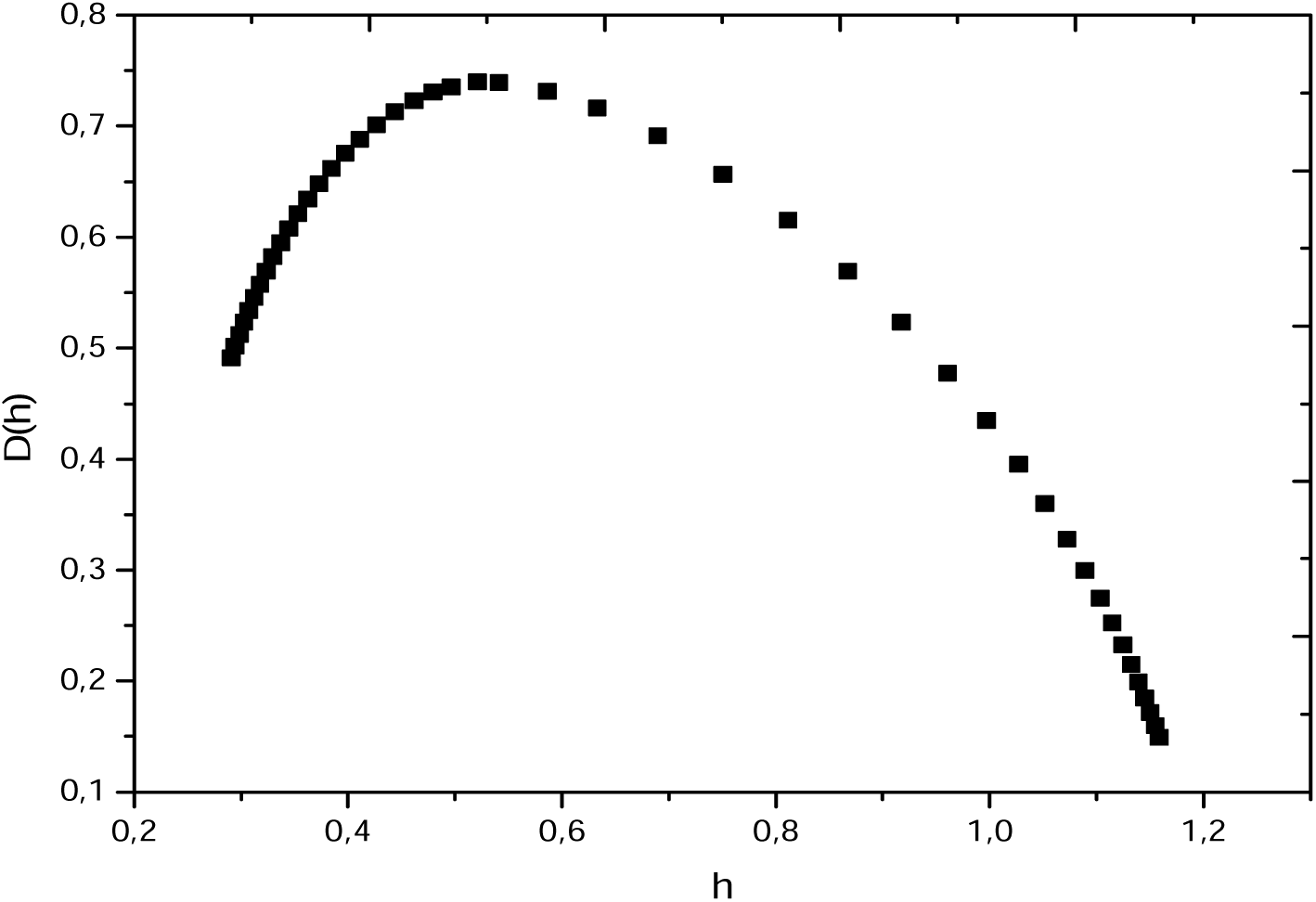
Spectrum of singularities of the daily death in Italy

## Conclusion

We have analyzed the daily and death cases of SARS-CoV-2 COVID-19 pandemic using the wavelet transform modulus maxima lines method, data of the World Health Organization are used. The case of Algeria, USA, Federation of Russia and Italy are processed, the multifractal behavior in all time series is demonstrated.

The dominance of the high or low Holder exponents in the spectrum of singularities of daily cases is controlled by the random respect of distancing and confinement protocols imposed by the governments, while the dominance in the daily death cases is mainly controlled by the health state and immunity of the infected patients (presence of chronic deseasses).

Decrease and low values of the daily death number in Italy is most probably due to the current version of SARS-CoV-2 coronavirus (which undergoes a lot of mutations) which currently less dangerous and fatal.

COVID-19 daily cases in Algeria fellow a randomized Gaussian model, while the daily death in Federation of Russia fellow the randomized Takagi model.

A second wave of the pandemic seems to be in the front in Italy by consequence a new protocol is imposed.

SARS-CoV2 COVID-19 pandemic spread in world fellows a variety of models, modeling them is a challenge, each model depends on the confinement, distancing and other protocols and respect of these protocols, traditions of peoples in term of malpractices regarding these protocols, architectures of the cities.

## Data Availability

Data are available via Health World Organisation dashbord
https://covid19.who.int/?gclid=CjwKCAjw74b7BRA_EiwAF8yHFBDbuMMyuNxuvJd_Z8Ut3h_ntiaVMqGfbFen4GekgwkWuGbtjvyBCRoCEGYQAvD_BwE

